# CT4CMS: Preoperative Computed Tomography-Based Consensus Molecular Subtyping Prediction in Colorectal Cancer Using Interpretable Deep Learning

**DOI:** 10.64898/2026.03.08.26347898

**Authors:** Xianrui Zhang, Xiuping Nie, Tan Wu, Du Cai, Hua Xue, Lin Qi, Yijuan Wang, Yi Cao, Lingli He, Yinghan Zhang, Yuanyao Cheng, Haotian Wang, Xiaoyu Wang, Enyu Li, Yue Dong, Feng Gao, Xin Wang

## Abstract

Consensus molecular subtyping (CMS) defines the transcriptomic taxonomy of colorectal cancer (CRC) and guides precision therapy. Although current approaches can predict CMS from histopathology, they rely on surgical specimens, limiting their preoperative applicability. In this study, we developed a deep learning model to infer CMS directly from preoperative computed tomography (CT) scans, enabling noninvasive molecular stratification of CRC. A multi-institutional cohort of 2,444 CRC patients was collected from the Sixth Affiliated Hospital of Sun Yat-sen University and Liaoning Cancer Hospital, comprising a discovery cohort (*n* = 416), an internal validation cohort (*n* = 1,671), and an external validation cohort (*n* = 357). To achieve robust feature extraction, a self-supervised 3D representation learning network was first pretrained on large-scale public CT datasets to capture generalizable imaging features. These representations were subsequently integrated into a multi-instance learning (MIL) classifier for CMS prediction, with attention mechanisms to enhance interpretability. Model performance was evaluated by cross-validation on the discovery cohort and verified on the two validation cohorts. CT4CMS demonstrated strong performance in predicting CMS subtypes directly from CT scans, achieving a cross-validation AUC of 0.867. In both validation cohorts, patients predicted as CMS4 exhibited significantly poorer disease-free survival yet derived substantial benefit from adjuvant chemotherapy, consistent with transcriptome-defined subtyping trends observed in the discovery cohort. Interpretability analysis revealed distinct subtype-specific radiomic features, suggesting that CT-derived imaging features capture underlying molecular characteristics and enable CMS classification. Overall, this study establishes a noninvasive and interpretable deep learning framework for CMS prediction in CRC, paving the way for imaging-based molecular stratification and personalized therapeutic decision-making.

## Introduction

Colorectal cancer (CRC) is the third most commonly diagnosed malignancy worldwide and exhibits the second fastest-growing incidence among all gastrointestinal cancers^1–3^. The disease presents substantial molecular heterogeneity, underscoring the importance of molecular subtyping for stratifying patients into more homogeneous groups, thereby enabling targeted treatment strategies and more accurate prognostic assessments^4–6^. Among current classification systems, consensus molecular subtyping (CMS) represents the most robust and widely accepted approach, offering significant prognostic and predictive values for CRC patients^7^. CMS subdivides CRC into four distinct molecular subtypes based on gene expression profiles, each associated with unique biological characteristics and clinical outcomes: CMS1 (microsatellite instability immune, favorable prognosis in early stages but poor outcomes in metastatic disease), CMS2 (canonical epithelial, intermediate prognosis), CMS3 (epithelial with metabolic dysregulation, intermediate prognosis), and CMS4 (mesenchymal with marked transforming growth factor-β activation, poor prognosis)^4–6^.

Currently, CMS classification relies primarily on next-generation RNA sequencing, a resource-intensive technique that poses significant challenges for widespread clinical adoption due to a high cost, lack of standardization, and the need for specialized infrastructure and bioinformatics expertise. These limitations restrict its availability to tertiary medical centers, leaving most local healthcare facilities unable to perform such testing^8^. To address these limitations, several studies have investigated hematoxylin and eosin (H&E)-stained histopathological slides as a cost-effective alternative for CMS prediction^5^. However, this approach depends on whole-slide images obtained after surgical resection, making it impractical for patients with inoperable tumors. Moreover, histology-based CMS assessment, typically performed postoperatively, cannot guide therapeutic strategies such as the use of neoadjuvant chemotherapy. Furthermore, conventional biopsies suffer from sampling bias and may not adequately represent the tumor’s molecular landscape. These challenges highlight the urgent need for a noninvasive, cost-effective, and robust approach to preoperatively identify CMS subtypes, thereby enhancing treatment stratification before surgery.

Computed tomography (CT), routinely performed as part of preoperative evaluation in CRC, offers a noninvasive imaging modality that has been widely adopted for diagnosis and prognostication. Radiomics, a burgeoning field in precision medicine, leverages high-throughput quantitative analysis of medical imaging data, particularly CT scans, to uncover patterns correlated with tumor biology^9^. In parallel, deep learning algorithms have demonstrated exceptional performance in analyzing CT images^10–13^, revealing meaningful associations between CT-derived radiomic signatures and molecular characteristics such as microsatellite instability (MSI) and *KRAS* mutations^14–16^. These advances suggested the potential to employ deep learning to predict CMS subtypes directly from preoperative CT images.

To the best of our knowledge, no prior research has demonstrated the feasibility of CMS prediction using deep learning features derived from CT images and elucidated the underlying imaging-molecular associations driving the model’s predictions. Furthermore, whether deep learning approaches outperform traditional handcrafted radiomics features in this task remains an open question. In this study, we propose a novel CT-based deep learning framework to preoperatively predict CMS subtypes in CRC patients. The main contributions of this work include: (1) the development of an interpretable deep learning model for CMS classification using pretreatment CT images; (2) the integration of a multiscale self-supervised learning strategy to enhance 3D feature extraction; (3) the implementation of an attention-based multi-instance learning (MIL) mechanism that enables accurate classification while preserving model interpretability; and (4) the introduction of a novel preoperative, CMS-informed decision point into the current clinical workflow to facillitate personalized treatment planning.

## Results

### Study design

The overall study design is illustrated in **Figure 1**. A total of 2,444 CRC cases from three independent cohorts were collected, comprising a discovery cohort SYSU-CRC (*n* = 416), an internal validation cohort SYSU-SAH (*n* = 1,671), and an external validation cohort Liaoning (*n* = 357) (**Figure 1A**). An interpretable deep learning framework, CT4CMS, was proposed to predict CMS in CRC patients using preoperative CT scans (**Figure 1B**). The framework integrated self-supervised pretraining and attention-based multi-instance learning (MIL) to achieve accurate and interpretable subtype classification. Model training and cross-validation were conducted in the discovery cohort SYSU-CRC, using paired CT images and corresponding gene expression data to provide ground-truth CMS labels for model development. The trained model was then independently tested in the two validation cohorts, SYSU-SAH and Liaoning, using only CT images for CMS prediction (**Figure 1C**). The predicted CMS subtypes were further evaluated for associations with patient survival outcomes, adjuvant chemotherapy benefit, and radiomics signatures, providing a comprehensive assessment of the model’s predictive performance and clinical relevance.

**Figure 1.**
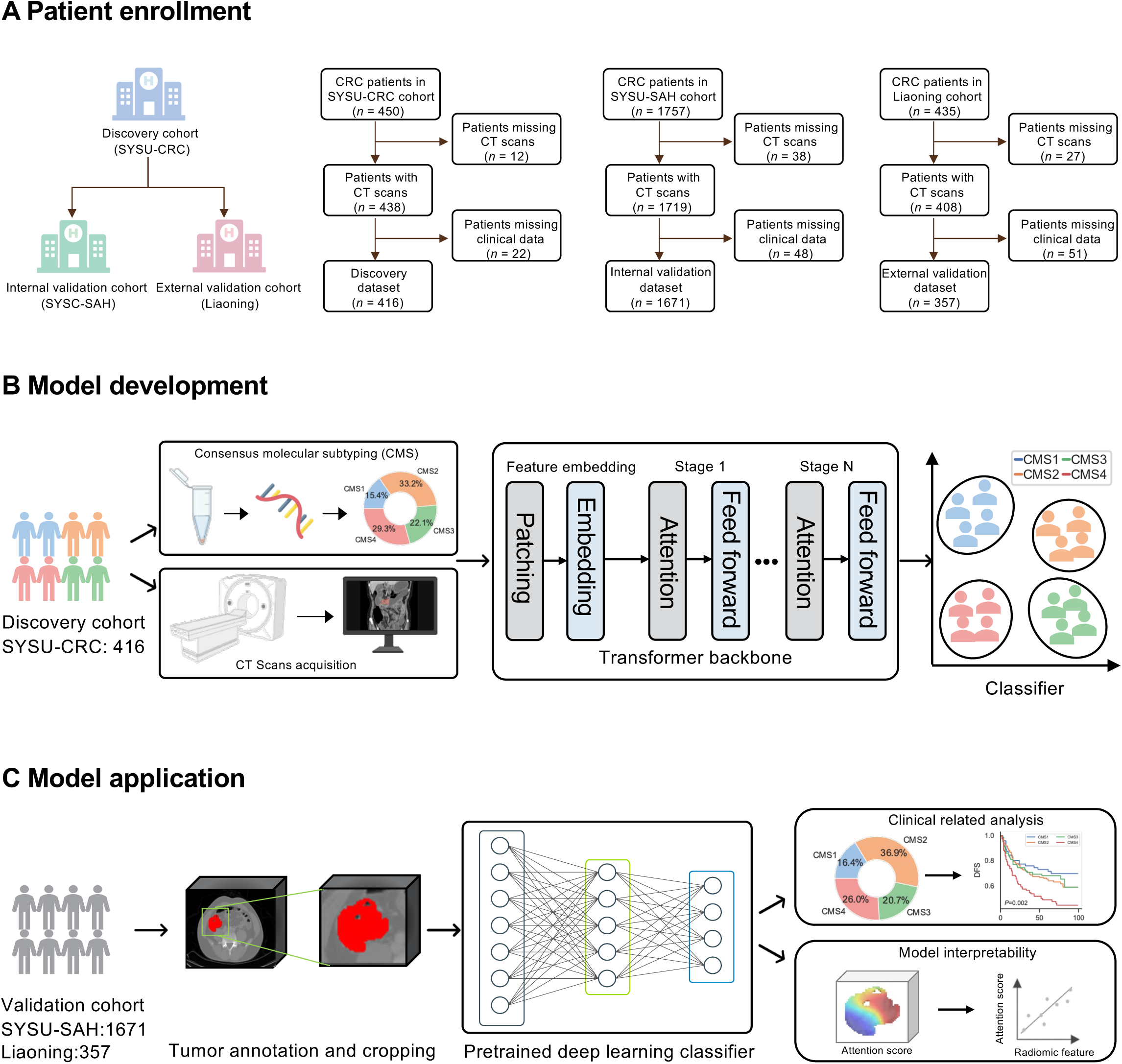
An overview of study design, model development, and application of the CT4CMS framework. **(A)** Patient enrollment across three independent centers, including the SYSU-CRC discovery cohort (*n* = 416), the SYSU-SAH internal validation cohort (*n* = 1,671), and the Liaoning external validation cohort (*n* = 357). In total, 2,444 patients with colorectal cancer (CRC) were retrospectively included, with those lacking preoperative CT scans or corresponding clinical data excluded. **(B)** Model development using paired preoperative CT scans and gene-expression profiles from the discovery cohort. Consensus molecular subtypes (CMS1-CMS4) derived from transcriptomic data served as ground-truth labels. A transformer-based deep learning architecture integrating patch embedding, hierarchical feature extraction, and attention-based multi-instance learning was trained to classify CRC into four CMS subtypes. **(C)** Model application and validation in independent cohorts. Annotated tumor regions were processed using the pretrained CT4CMS classifier to generate CMS predictions. The predicted CMS subtypes were further analyzed for clinical relevance, including associations with survival outcomes, chemotherapy response, and model interpretability assessed by attention heatmaps and radiomic feature correlations.

### CT4CMS integrated self-supervised learning and attention-based aggregation for interpretable CMS prediction

The CT4CMS framework comprises four main stages (**Figure 2**). First, tumor regions within 3D CT volumes were segmented (**Figure 2A**). Second, a Swin-Transformer backbone pretrained with a multiscale masked image modeling (MIM) strategy^17^ captured multiscale contextual information from medical images (**Figure 2B**). This self-supervised pretraining enabled the model to learn high-level morphology and texture patterns across diverse CT domains. Third, the pretrained backbone generated 3D feature instances that were aggregated by an attention-based multi-instance learning (MIL) classifier to predict CMS subtypes (**Figure 2C**). Ground-truth CMS labels in the discovery cohort were derived from gene-expression profiles using the established pipeline described by Wang et al.^6^. Finally, attention scores from the MIL classifier provided interpretability by highlighting the image regions most relevant to the model’s predictions and linking them to radiomic features (**Figure 2D**). These radiomic features were subsequently analyzed and correlated with predicted CMS subtypes, demonstrating that CT4CMS captured biologically and clinically relevant phenotypes.

**Figure 2.**
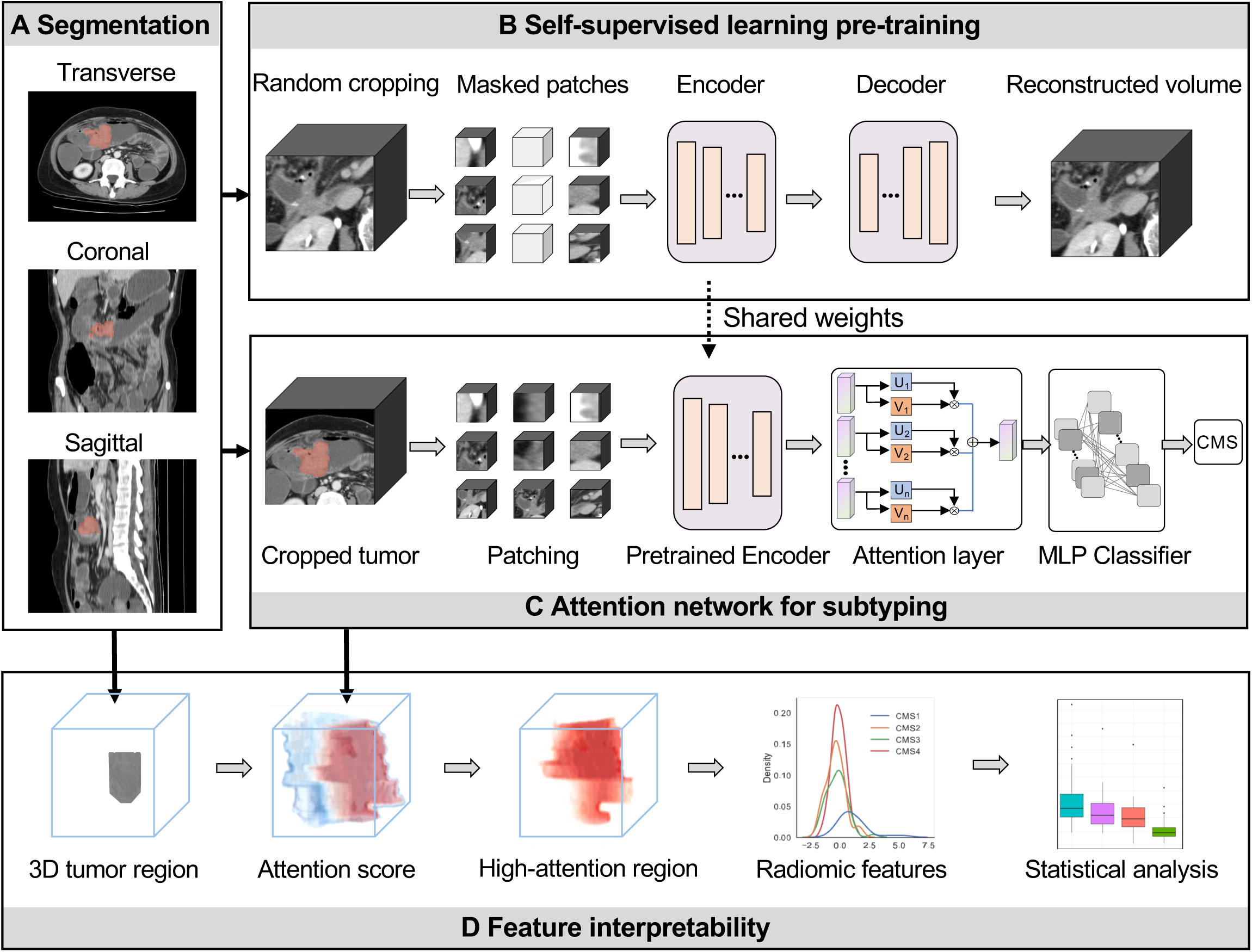
The overall framework of the CT4CMS model. **(A)** Tumor regions were manually annotated from computed tomography (CT) images by experienced radiologists. **(B)** Self-supervised pretraining for representation learning using masked image modeling. Three-dimensional image volumes (96 × 96 × 96 voxels) were randomly cropped from whole CT scans to construct a training dataset for masked patch reconstruction. **(C)** Downstream model for consensus molecular subtyping (CMS) classification. Deep learning features were extracted from cropped tumor volumes using a pretrained Swin-Transformer encoder. The final-layer feature maps were aggregated via an attention layer, and a multilayer perceptron (MLP) classifier was employed to predict CMS subtypes. **(D)** Model interpretability and radiomic correlation analysis. Attention heatmaps were generated to highlight image regions contributing to CMS prediction, with blue indicating low-attention and red indicating high-attention areas. Radiomic features, including first-order statistics, second-order morphological and high-level texture descriptors, were extracted from high-attention regions and analyzed for associations with molecular subtypes.

### CT4CMS accurately predicted CMS subtypes of CRC

Model performance was first assessed using five-fold cross-validation on the SYSU-CRC cohort. CT4CMS achieved strong classification performance across all CMS subtypes (**Figure 3A-D**). The ROC comparison demonstrated that CT4CMS achieved the highest AUC of 0.867, outperforming three benchmark models: a radiomics-based model (AUC = 0.586), SwinUnetr^18^ (AUC = 0.736), and MaskedIM^19^ (AUC = 0.807) (**Figure 3A**). More specifically, the radiomics-based model, which relied on 816 handcrafted features and an MLP classifier, showed limited discriminative ability. By contrast, both SwinUnetr and MaskedIM leveraged self-supervised feature extraction with the same MIL classifier and exhibited better performance, yet still fell short of CT4CMS. Moreover, CT4CMS consistently surpassed all baselines in accuracy, precision and recall, confirming the superior representation power of its multiscale self-supervised learning strategy for 3D CT data (**Table S2**).

**Figure 3.**
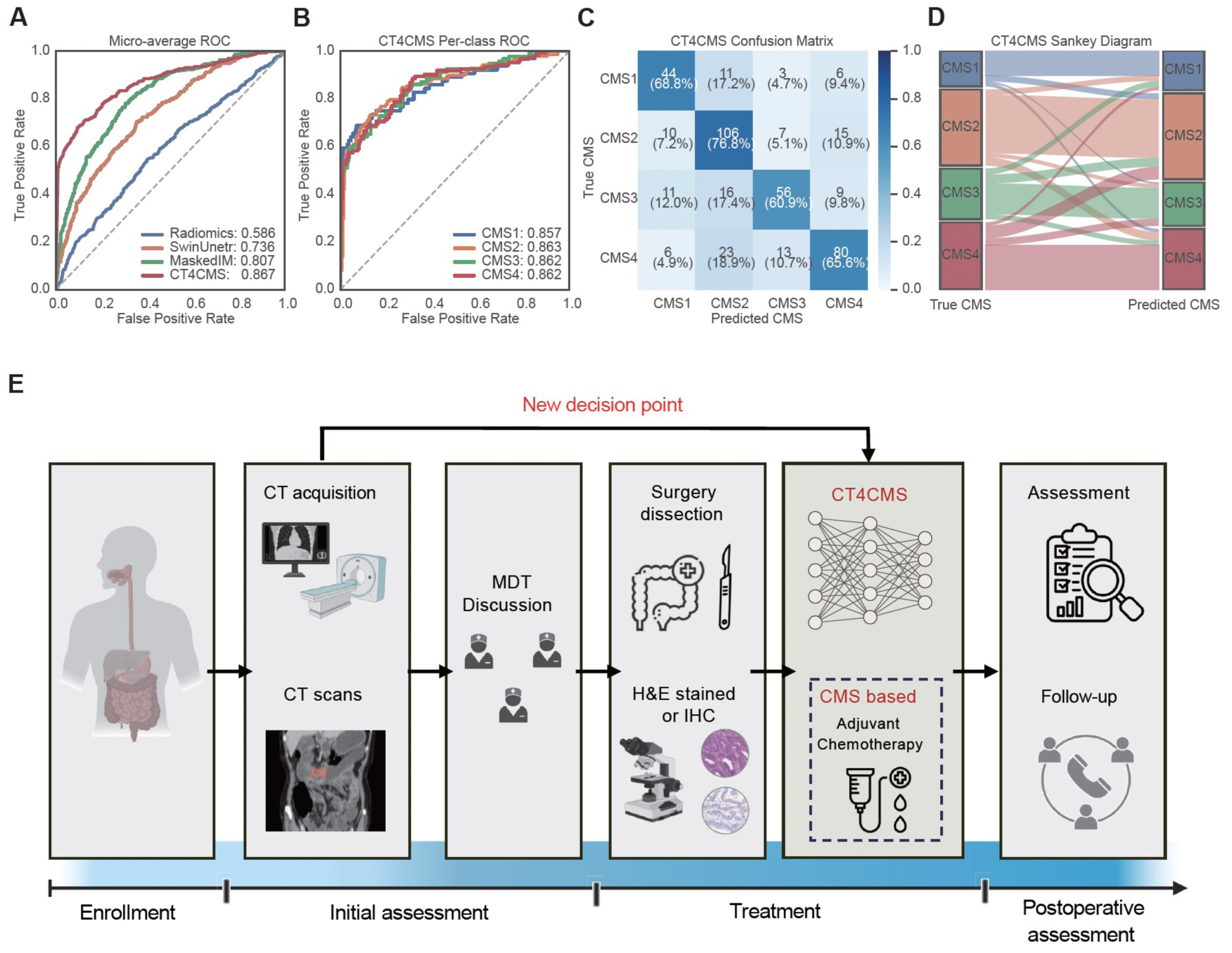
High performance of CT4CMS enables a new preoperative decision point in the clinical workflow. **(A)** Receiver operating characteristic (ROC) curves comparing CT4CMS with benchmark models, demonstrating superior discrimination performance (AUC = 0.867). **(B)** Per-subtype ROC curves confirming balanced predictive performance across all four consensus molecular subtypes. **(C)** Confusion matrix showing accurate classification. **(D)** Sankey diagram illustrating strong alignment between true and predicted CMS labels. **(E)** Integration of CT4CMS into the clinical workflow enables preoperative, noninvasive CMS prediction from CT scans to assist multidisciplinary team (MDT) discussions, guide adjuvant chemotherapy and support postoperative follow-up.

The per-subtype ROC curves further indicate that CT4CMS maintained stable and balanced classification performance, with AUCs of 0.857, 0.863, 0.862, and 0.862 for CMS1 to 4, respectively (**Figure 3B**). The confusion matrix shows that most predictions aligned with their true CMS labels, with particularly accurate recognition of CMS4, a clinically important subtype associated with poor prognosis (80 correctly classified cases) (**Figure 3C**). The Sankey diagram illustrates the correspondence between true and predicted labels, where the strong diagonal flows confirm the model’s subtype-specific consistency with minimal cross-category confusion (**Figure 3D**).

Based on the promising model performance, we propose to integrate CT4CMS into the current clinical workflow for non-invasive prediction of CRC subtypes to facilitate personalized treatment decisions (**Figure 3E**). Focusing on stage II/III CRC in this study, we hypothesize that the prediction of CMS preoperatively based on baseline CT scans can assist more accurate selection of patients for adjuvant chemotherapies after surgery.

### CT4CMS demonstrated consistent prognostic power across independent cohorts

To test our hypothesis, we first evaluated the prognostic value of CT4CMS-based molecular subtypes in three independent clinical cohorts. More specifically, Kaplan-Meier (KM) survival analyses were performed using disease-free survival (DFS) as the endpoint across the SYSU-CRC, SYSU-SAH, and Liaoning cohorts. In the SYSU-CRC discovery cohort, where transcriptomic CMS labels were available, CMS4 patients had significantly worse DFS than CMS1-3 (*P* = 0.025) (**Figure 4A**), confirming the strong prognostic relevance of transcriptome-defined stratification. In the two validation cohorts without gene-expression data, CT4CMS-predicted CMS labels reproduced the same pattern, with CMS4 again exhibiting the poorest DFS (SYSU-SAH, *P* = 3.69 × 10⁻⁶; Liaoning, *P* = 0.043) (**Figure 4B-C**). Cox regression analyses focusing on CMS subtypes further quantified these observations, demonstrating that CMS4 patients consistently had a higher risk of recurrence than other subtypes across all datasets (**Figure 4D**). To assess whether CMS4 retained independent prognostic value, univariable Cox regression analyses were first performed to identify factors associated with survival, and significant variables were subsequently included in multivariable Cox regression analyses, including CT4CMS predictions and conventional clinicopathologic variables (e.g., stage, sex, age, and grade), which confirmed CMS4 as an independent prognostic marker (**Figures S1-S6**). These findings validate CT4CMS as a robust, non-invasive method for prognostic stratification, faithfully reproducing transcriptomic CMS patterns using only preoperative CT scans.

**Figure 4.**
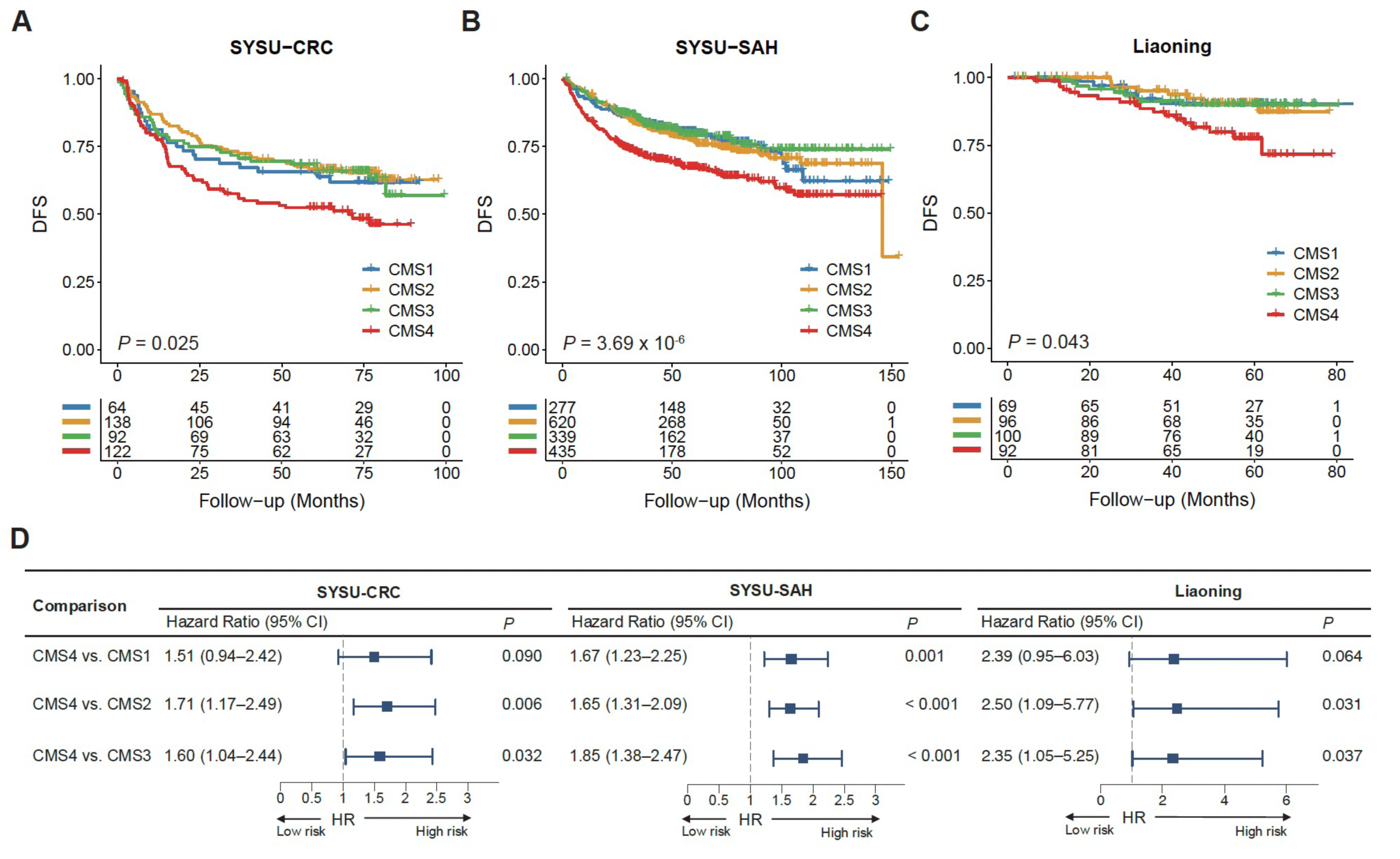
Prognostic value of CT4CMS-predicted consensus molecular subtypes across cohorts. **(A)** Kaplan-Meier curves of disease-free survival (DFS) in the SYSU-CRC cohort using transcriptomic CMS labels, showing significantly poorer outcomes for CMS4 patients (*P* = 0.025). **(B)** DFS curves in the SYSU-SAH validation cohort using CT4CMS-predicted CMS labels, with CMS4 again demonstrating the worst prognosis (*P* = 3.69 × 10⁻^6^). **(C)** DFS analysis in the Liaoning cohort (*P* = 0.043), confirming that CT4CMS-derived CMS predictions stratify patients into prognostically distinct subgroups consistent with transcriptomics-based classification. **(D)** Forest plots showing hazard ratios (HR) for pairwise comparisons between CMS4 and other subtypes across cohorts. CT4CMS-predicted CMS4 was consistently associated with an increased risk of recurrence, aligning with gene-expression-derived CMS characteristics.

### CT4CMS-predicted subtypes showed significant association with chemotherapy benefit

We next investigated the association between CT4CMS-predicted subtypes and the benefit of adjuvant chemotherapy. Across the SYSU-CRC, SYSU-SAH, and Liaoning cohorts, DFS was compared between surgery-only and adjuvant-treated patients within each CMS subtype. In the SYSU-CRC cohort, where ground-truth CMS labels were available, surgery-only patients showed clear survival differences across subtypes, with CMS4 having the poorest DFS (*P* = 3.46 × 10⁻⁴) (**Figure 5A**). Among patients who received adjuvant chemotherapy, this disparity disappeared (*P* = 0.92) (**Figure 5B**). Notably, stage II-III patients assigned to CMS4 who underwent adjuvant therapy significantly improved DFS compared with those treated by surgery alone (*P* = 3.68 × 10⁻⁵) (**Figure 5C**). The SYSU-SAH and Liaoning validation cohorts, using CT4CMS-predicted CMS labels, exhibited concordant patterns (**Figure 5D-I**). In both cohorts, CMS4 patients showed substantially poorer DFS following surgery alone (SYSU-SAH, *P* = 6.90 × 10⁻⁶; Liaoning, *P* = 3.49 × 10⁻³), but experienced significant improvement with adjuvant chemotherapy (SYSU-SAH, *P* = 1.36 × 10⁻⁴; Liaoning, *P* = 0.016). By contrast, CMS1-3 subtypes showed no appreciable benefit from chemotherapy (all *P* > 0.05, **Figure 5**). Collectively, these results indicate that adjuvant chemotherapy can mitigate the otherwise poor prognosis of the CMS4 subtype. The concordant patterns observed across both transcriptomic and CT-derived classifications underscore the clinical value of CT4CMS in identifying patients most likely to benefit from adjuvant chemotherapy, particularly in settings where genomic testing is unavailable.

**Figure 5.**
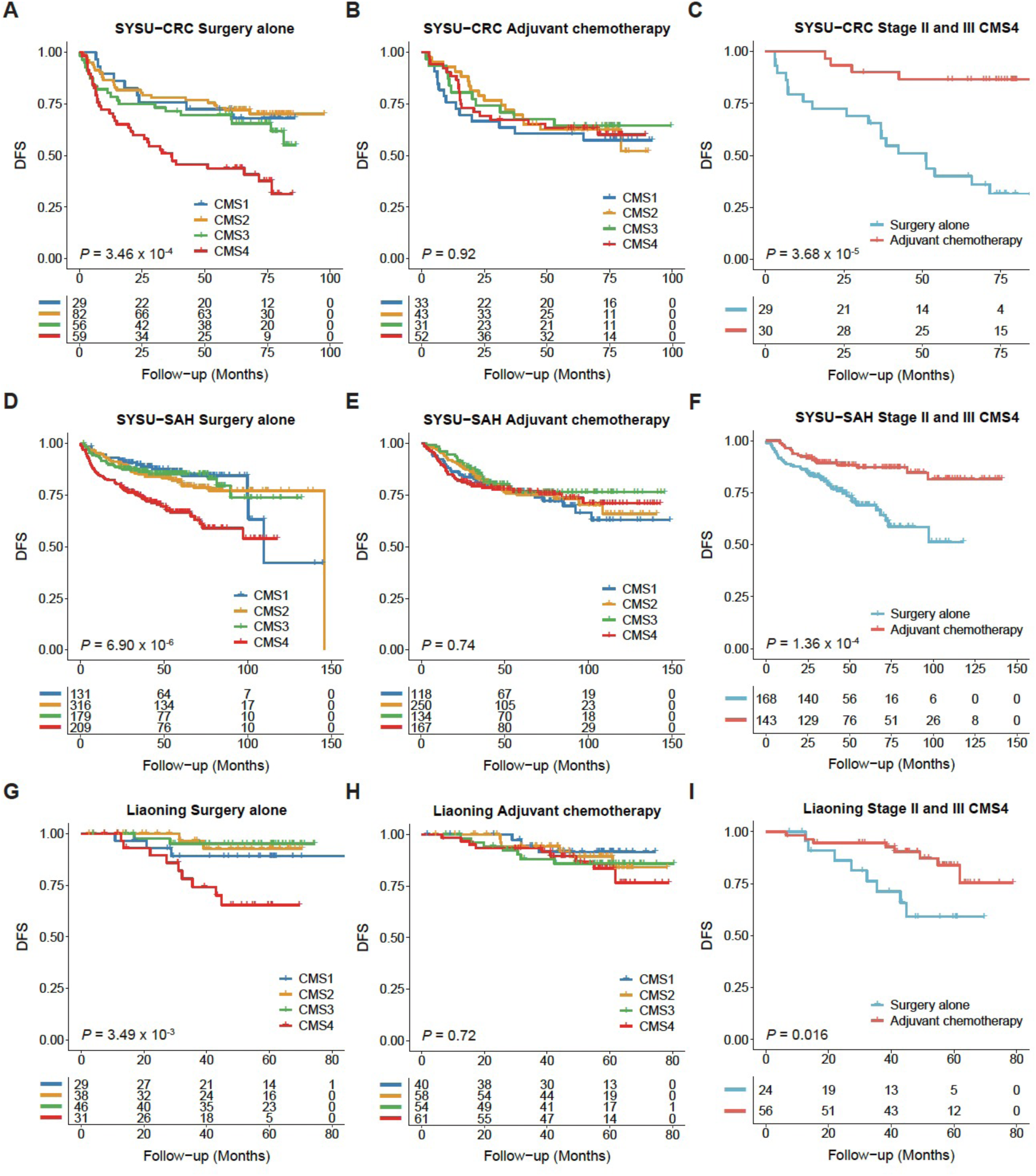
Association between CT4CMS-predicted consensus molecular subtypes and adjuvant chemotherapy benefit in colorectal cancer. (A-C) Kaplan-Meier curves of disease-free survival (DFS) in the SYSU-CRC cohort using transcriptomic CMS labels. CMS4 patients exhibited significantly poorer outcomes after surgery alone (*P* = 3.46 × 10⁻⁴), whereas adjuvant chemotherapy mitigated this survival disadvantage (*P* = 0.92). Among stage II and III CMS4 patients, those receiving adjuvant chemotherapy achieved markedly prolonged DFS (*P* = 3.68 × 10⁻⁵). **(D-F)** Validation in the SYSU-SAH cohort using CT4CMS-predicted CMS labels showed consistent results. Surgery-only CMS4 patients had inferior survival (*P* = 6.90 × 10⁻⁶), while adjuvant chemotherapy significantly improved outcomes among stage II and III CMS4 cases (*P* = 1.36 × 10⁻⁴). **(G-I)** Analyses in the Liaoning cohort confirmed similar trends. CT4CMS-predicted CMS4 patients again demonstrated the poorest DFS after surgery alone (*P* = 3.49 × 10⁻³), whereas adjuvant therapy conferred clear benefit in stage II and III CMS4 patients (*P* = 016).

### CT4CMS-predicted CMS4 patients benefited from adjuvant chemotherapy in stage II and III disease

To further evaluate the stage-specific implications of CT4CMS predictions, we analyzed adjuvant chemotherapy outcomes within stage II and III CRC patients across SYSU-CRC, SYSU-SAH, and Liaoning cohorts. Kaplan-Meier survival analyses stratified by treatment (surgery alone vs. adjuvant chemotherapy) revealed that only the CMS4 subgroup derived a significant benefit from adjuvant chemotherapy, whereas other CMS patients exhibited no improvement in DFS (**Figure S7-S9)**. In stage II disease, this pattern provides a potential solution to resolve the persistent clinical ambiguity regarding which patients should receive adjuvant therapy, with CMS4 tumors representing the only group with clear benefit (**Figure S8**). In stage III disease, although adjuvant chemotherapy is generally recommended for all patients, only the CMS4 group showed significant survival improvement, while non-CMS4 subtypes displayed no improvement or even unfavorable outcomes (**Figure S9**). Univariable Cox regression analyses across the three cohorts further supported these observations (**Figure S10-S12**). CMS4 patients receiving adjuvant chemotherapy had consistently lower risks of recurrence, with hazard ratios below one in all datasets, whereas CMS1-3 showed no significant improvement. Collectively, these results highlight that CT4CMS enables preoperative identification of chemo-sensitive CMS4 patients in both stage II and III disease. By distinguishing those truly likely to benefit from adjuvant chemotherapy, CT4CMS supports precision oncology and guides optimal treatment selection while avoiding unnecessary exposure to ineffective chemotherapy.

### Interpretability analysis revealed CMS-specific radiomics signatures at high-attention regions

To interpret the model’s high-attention regions, we quantified their imaging characteristics by extracting radiomic features and analyzing their associations with attention scores (**Figure 2D**). A total of 851 radiomic features were extracted from these regions, including first-order statistical features, second-order morphological features, and texture-based descriptors^20,21^. Statistical analyses using the Wilcoxon rank-sum test and analysis of variance (ANOVA) identified 51 features that showed statistically significant differences across CMS subtypes. These selected radiomics features revealed clear subtype-specific associations (**Figure 6**), suggesting that the regions highlighted by CT4CMS captured distinctive structural and textural phenotypes of each molecular subtype. For example, CMS1 tumors showed higher values in Grey Level Emphasis (GLE) related features, indicating less uniformity in CT images, which is consistent with elevated immune cell infiltration (*P* < 0.001). CMS2 tumors exhibited features of Skewness and 90 Percentile, which could be associated with epithelial differentiation and a high density of tumor cells (*P* < 0.001). CMS3 tumors were characterized by Long Run Emphasis and larger two-dimensional diameters in axial slices, reflecting increased mucus content (*P* < 0.001). CMS4 tumors displayed features such as Dependency Entropy, Variance, and Cluster Tendency, indicating greater heterogeneity, which aligns with the presence of stromal invasion and angiogenesis observed in our previous studies (*P* < 0.001). Collectively, these findings demonstrate that CT-derived imaging features can reflect the underlying molecular characteristics of CRC and thereby enable accurate CMS classification.

**Figure 6.**
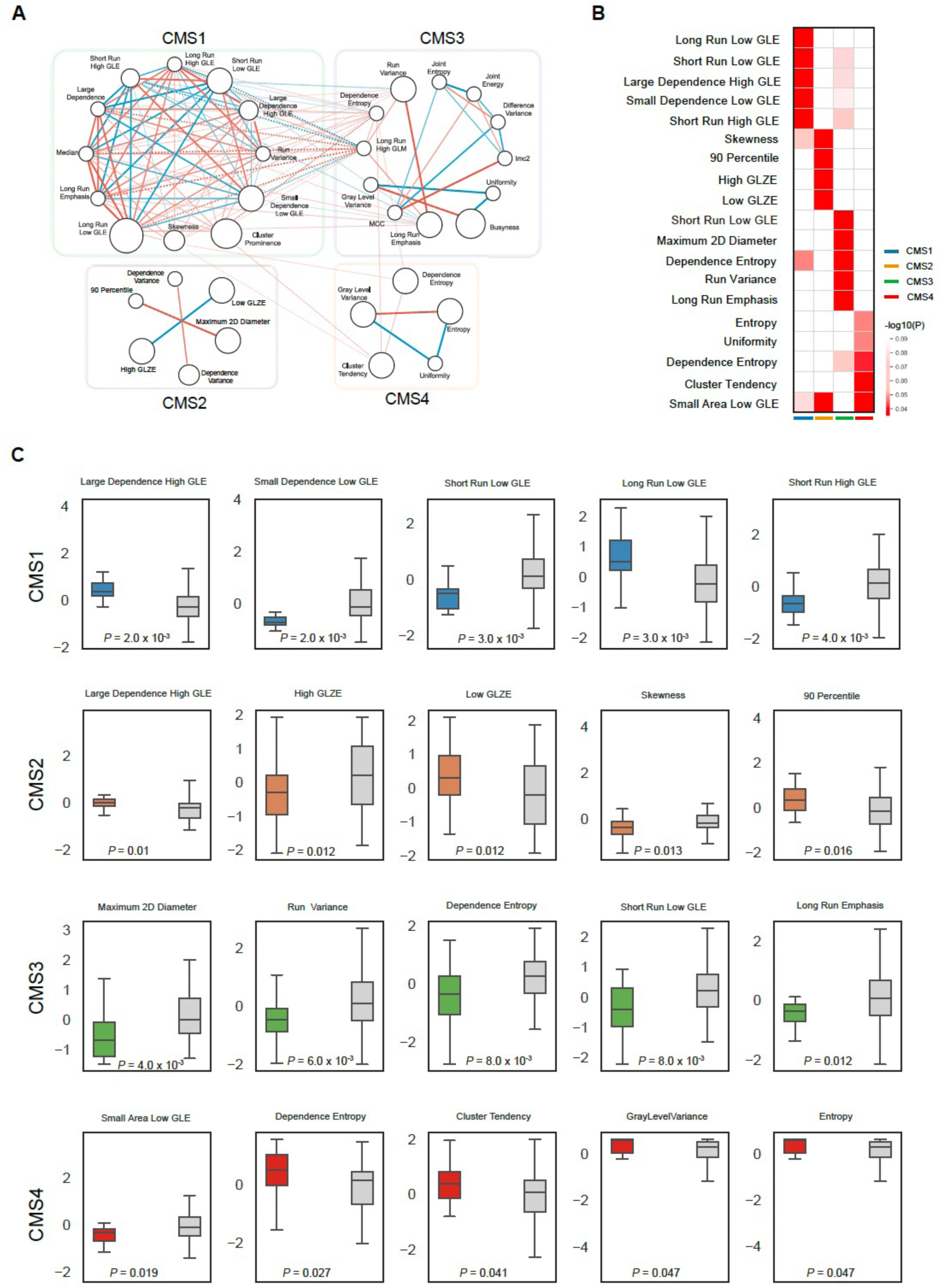
Radiomic interpretation of CT4CMS-predicted molecular subtypes. **(A)** Network analysis of the most significant radiomic features extracted from high-attention regions for each consensus molecular subtype (CMS), revealing subtype-specific feature interactions. **(B)** Heatmap displaying the −log₁₀(*P*) values from Wilcoxon rank-sum tests, highlighting radiomic features significantly associated with individual CMS subtypes. **(C)** Comparison of representative radiomic features across CMS groups in the discovery cohort. For each subtype, the five most discriminative features are shown, comparing feature value distributions between that subtype and all other subtypes. These results demonstrate distinct imaging phenotypes underlying CT4CMS-predicted consensus molecular subtypes.

## Discussion

In this study, we developed a deep learning framework, CT4CMS, that enables noninvasive, preoperative prediction of CMS using routine CT imaging. The model was trained on a cohort of 416 patients with paired imaging and transcriptomic data and validated on two independent cohorts totaling 2,028 patients. CT4CMS demonstrated robust classification performance across all CMS groups and provided interpretable outputs that support clinical translation.

Previous radiomics studies have primarily focused on tumor staging, lymph node assessment, or MSI prediction, with few attempts at CMS classification using contrast-enhanced CT. Although RNA sequencing remains the gold standard for CMS determination, it is invasive, costly, and not always feasible in routine clinical practice. In contrast, CT imaging offers a noninvasive, repeatable, and whole-tumor assessment method, making it an ideal modality for molecular stratification.

To address the limitations of existing approaches, we introduced a multiscale masked image model for self-supervised pretraining. This strategy enhanced feature representation and model generalizability, outperforming both radiomics-based classifiers and other self-supervised learning methods. The proposed framework achieved an AUC of 0.867 in the internal cohort and showed consistent performance across external datasets, confirming its robustness. By integrating attention scores with radiomic profiling, CT4CMS provided interpretable outputs that revealed distinct patterns for each CMS subtype, consistent with known histopathological and molecular characteristics.

Importantly, CT4CMS also demonstrated predictive value in therapeutic decision-making. Across three cohorts, patients with predicted CMS4 consistently showed poor survival outcomes but derived significant benefit from adjuvant chemotherapy. In contrast, no clear benefit was observed in CMS1, CMS2, or CMS3 patients. These findings suggest that CT4CMS can help identify high-risk patients who may require more intensive treatment and spare others from unnecessary therapy.

The current clinical workflow for CRC involves an initial clinical assessment, including medical history, physical examination, and screening tests such as fecal occult blood test, carbohydrate antigen 19-9 (CA 19-9), and carcinoembryonic antigen (CEA), followed by diagnostic confirmation through colonoscopy with tissue biopsy. Imaging examinations such as CT, MRI, and PET are then used for disease staging. Preoperative evaluation generally includes laboratory testing and multidisciplinary team (MDT) consultations to establish individualized treatment strategies. Within this workflow, CT4CMS can be integrated after CT acquisition to provide noninvasive, preoperative prediction of CMS subtypes directly from routine CT images. This integration augments conventional anatomic and pathological information with molecular insights, enabling CMS-based treatment planning before surgery. As illustrated in **Figure 3E**, CT4CMS-generated predictions can assist MDT discussions and guide personalized clinical decisions.

This study has several limitations. First, its retrospective design and potential selection bias may affect generalizability. We mitigated this by including large, multi-center cohorts and adjusting for clinicopathological variables in survival analyses. Second, due to the irregular shape and low contrast of CRC tumors on CT, manual segmentation was required for accurate ROI extraction, introducing variability. Current segmentation algorithms remain insufficiently reliable for fully automated contouring in colorectal CT. Future work should focus on developing advanced methods for automated or semi-automated tumor segmentation to facilitate clinical implementation.

In conclusion, CT4CMS provides a clinically meaningful and interpretable deep learning approach for noninvasive CMS prediction using routine preoperative CT imaging. By enabling molecular subtyping, treatment guidance, and prognosis estimation without the need for invasive procedures, this model advances precision oncology in CRC. While further validation in larger prospective cohorts is needed, this study establishes a foundation for future radiomics and AI research in CRC and beyond.

## Materials and methods

### Patient cohorts and data collection

In this study, we collected five publicly available CT datasets comprising more than 5,000 abdominal CT volumes, including AbdomenCT-1K^22^, BTCV^23^, MSD^3^, WORD^24^, and TCIA-Colon^25^. In addition, three clinical cohorts were retrospectively collected, including the discovery cohort (SUSY-CRC) and the internal validation cohort (SYSU-SAH) from the Six Affiliated Hospital of Sun Yat-sen University, and the external validation cohort (Liaoning) from Liaoning Cancer Hospital. The SYSU-CRC cohort included 416 consecutive patients with both CT images and gene expression profiles. The SYSU-SAH and Liaoning cohorts consisted of 1,671 and 357 patients, respectively, each providing only CT imaging data. Clinicopathological characteristics of all included patients are detailed in **Table S1**. Ethical approval was obtained from the medical ethics committees of Sun Yat-sen University and Liaoning Cancer Hospital, and informed consent was obtained.

Inclusion criteria were as follows: (i) histopathologically confirmed primary CRC; (ii) availability of contrast-enhanced abdominopelvic CT scans performed within two weeks prior to surgery. Patients were excluded if they had (i) incomplete clinicopathological information; (ii) a time interval exceeding two weeks between CT examination and surgery; or (iii) inoperability or refusal of surgical treatment. Baseline clinical data, including age, sex, TNM stage, grade, microsatellite instability (MSI) status, chemotherapy treatment, and disease-free survival (DFS), were collected for all patients (**Tables S3-S5**).

### CT image preprocessing

All raw CT scans from public datasets were preprocessed and used in self-supervised learning. The SYSU-CRC cohort was trained and validated by the CMS classifier through 5-fold cross-validation. The ground truth label of CMS is obtained from gene expression profiles of corresponding patients. The SYSU-SAH and Liaoning cohorts were used as independent validation cohorts. Due to the lack of gene expression data and CMS information, they were employed as independent test sets to evaluate model performance based on correlation with CMS types via survival analysis. For pretraining the feature extractor and training the classifier, CT images were resampled to a pixel space of 1.5 mm in-plane and 3 mm for axial slice thickness. HU values were clipped to −1000 to 1000 and normalized to 0 to 1. Only raw CT images were used in the self-supervised learning stage. The MIL classifier was trained using tumor region annotations. Two radiologists from SYSU and Liaoning, each with more than five years of experience in abdominal imaging, manually delineated the tumor regions on preoperative CT scans. The annotations were reviewed independently by both readers, and any discrepancies were resolved by consensus after joint discussion.

### Development of a multi-scale self-supervised model

The overall architecture of CT4CMS was optimized using a multi-scale self-supervised learning strategy (**Figure 2**). The 3D Swin-Transformer^26^, employed as the backbone of the feature extractor, directly utilizes 3D patches and is the most advanced architecture. It has been used in many medical image-related studies, such as Swin-UNETR, Disruptive Autoencoder, and SwinMM^18,19,27^. In this study, it was reimplemented to 3D versions to accommodate the 3D volume data of CT scans and was used as the backbone for both pretraining and downstream classification tasks. The details of the self-supervised learning using a multiscale masked imaging model (MIM) are illustrated in **Figure S13**.

The patch embedding layer transforms data into sequences. Original 3D volumes *X*, with the shape of *C × D × H × W*, are reshaped into sequences of *N* regular non-overlapping small 3D patches, where *C* = 1 is the number of channels, and *D, W* and *H* represent the depth, width, and height of a 3D image. A trainable linear projection maps these patches to embeddings. For instance, a 3D volume with a size of 1 × 96 × 96 × 96 is divided into 216 smaller patches of 16 × 16 × 16 or 110,592 smaller patches of 2 × 2 × 2, considered the basic processing units of the encoder. A uniformly random masking method at the patch level with a high mask ratio effectively eliminates redundancy, creating a self-supervisory task that cannot be easily solved by extrapolating from visible neighboring patches^18^. Meanwhile, the uniform distribution avoids potential center bias (i.e., more masked patches near the image center). Additionally, since the number of patches *N* decreases through layers in the pyramidal architectures of Swin-Transformer, when constructing the random mask for inputs, we first mask the image randomly with the size of the last stage’s input in Swin-Transformer and then up-sample it to the same size as the inputs^17^. Next, the Swin-Transformer takes the visible patch embeddings and learnable mask embeddings as inputs and is responsible for modeling latent feature representations of the masked patches. Therefore, the original image resolution was down-sampled in different stages with the scale of 1/4,1/8,1/16 and 1/16.

Based on the outputs in different stages from the previous Swin-Transformer encoder, the latent features in different scales will be utilized to forecast the original signals in the masked area in the decoders. In each stage, the corresponding decoder takes the entire encoded token collection, including visible patches and mask tokens, as inputs. Each randomly initialized mask token is a learnable vector jointly optimized to reveal the masked patches. The absolute positional embeddings are also applied to these mask tokens corresponding to the backbone architecture^17^. Then, we thoroughly investigate the effectiveness of MIM models in different scales and reconstruct the inputs by estimating the raw voxel values for each mask token simply and intuitively from 3D medical imaging data. We use tiny decoders containing one transformer block with a small embedding dimension and one multilayer perceptron layer to predict pixel values (**Figure S13**). It has been approved that a lightweight decoder avoids excessive computational overhead and increases the encoder’s ability to learn more generalizable representations^28,29^. Also, in our study, the encoder’s well-optimization is more important as only the encoder would be inherited for downstream learning.

Reconstruction strategies for images by decoders on different scales ensure lower layers learn low-level information and upper layers learn high-level information^17^. Thus, it is not appropriate to use single-scale supervision to guide multiple local layers based on the final outputs of the Swin-Transformer. The finely divided input regions typically contain the low-level semantical information of the input, like corners, edges, or textures. Relatively, the coarse-scale supervisions capture high-level semantical information of the input, like the shape of the partial or whole object. Intuitively, multiscale supervisions can guide representation learning better than the common single-scale ones due to their richer semantic information. To this end, we make the lower layers to reconstruct fine-scale supervisions and the upper ones to reconstruct coarse-scale supervisions for multiscale reconstruction of normalized pixels. For compatibility, we use the supervisions with the same scale as the feature maps from different stages in Swin-Transformer, as its pyramidal architecture already hardcodes the multiscale property to the features by setting their spatial sizes.

During pretraining, we randomly flip and crop a 1 × 96 × 96 × 96 volume as the input for Swin-Transformer. The initial learning rate is 1.5 × 10^−4^ following a cosine decay schedule, and weight decay is 0.05. The multiscale MIM pretraining runs for 300 epochs on all the datasets with batch sizes of 2. The training loss is the weighted summation of the reconstruction losses at the chosen layers and guides the patches at multiple chosen layers to conduct semantic interactions under different scales, which not only accelerates the learning of multiple layers but also facilitates multiscale semantical understanding of the input. Finally, we optimized the model by minimizing the distance between recovered and original images in voxel space using *L2* loss computed on masked patches only. It could prevent the model from engaging in self-reconstruction, which might potentially dominate the learning process and ultimately impede knowledge learning^17,18,28,29^.

We showed the reconstruction results of multiscale MIM with a mask ratio of 75% (**Figure S14**). The three rows show the original images, the masked images, and the reconstructed images, respectively. The results demonstrated that multiscale MIM could restore the lost information from the random context. It is worth noting that MIM aims to benefit downstream tasks rather than generating high-quality reconstructions. We hypothesize that the contextual information for reconstructions of masked image patches is particularly important in medical imaging tasks where any given ROI is intrinsically dependent and connected to its physiological environment and surroundings.

### Development of the CMS classifier by multi-instance learning

After feeding the original CT image with shape *C × D × H × W*, the feature extractor Swin-Transformer generates abstract features with shape *C_1_ × D_1_ × H_1_ × W_1_* (**Figure 2**). Based on the flattened features, we will obtain a bag of deep 3D instances *H,* containing *D_1_ × H_1_ × W_1_* instances with *C_1_* channels. Then we embedded the attention-based MIL pooling approach into the framework for global feature aggregation. Finally, a multilayer perceptron was trained for CMS prediction using aggregated features.

The main difference from the existing attention mechanism is that we apply it to the 3D instance feature *H*. Intuitively, if a deep 3D instance receives the largest attention weight, it is the key instance. The attention weights can give insight into every instance’s contribution to the bag label. Therefore, attention-based MIL pooling gives strong interpretability to predictions. Also, the generated bag representation from attention-based MIL is more semantic than traditional MIL pooling operators. Therefore, based on a pretrained 3D Swin-Transformer from multiscale MIM, the attention-based MIL pooling module can receive deep 3D instances and generate the semantic representation for 3D CT images compared with the simple 2D slicer-based CT model.

In MIL training, the Swin-Transformer parameters were frozen and used as the feature extractor. The learning rate of MIL is set to 1 × 10^−3^ and the batch size is 1. The learning rate during training MIL also follows a cosine decay schedule. Receiver operating characteristic curve (ROC) analysis was performed to evaluate the performance of MIL, and then the area under the receiver operating characteristic curve (AUC), sensitivity, specificity, and F1 score were calculated in the validation cohort.

### Interpreting the deep learning model via radiomic features and attention scores

Attention scores from the MIL model, before conversion to a probability distribution by the softmax function, are computed for all patches extracted from the tumor region using the attention branch corresponding to the predicted class. Converted to percentile scores and scaled between 0 and 1, patches within tumor regions with attention scores greater than 0.5 are selected as high-attention regions. Normalized scores are converted to RGB colors using a diverging colormap, displaying high attention in red and low attention in blue.

After creating attention maps, radiomic profiling is applied to extract features from the selected highly attended region in contrast-enhanced CT scans. Image filters with multiple scales were applied to original CT images, including the Laplacian of Gaussian (LoG) filter with sigma 1.0, 2.0 and 3.0 mm and wavelet filter^21^. Together, the workflow produces 851 features for each CT scan. Using the workflow, we performed radiomics profiling for CT images in the discovery cohort SYSU-CRC, and 851 radiomics features were extracted. Next, the Wilcoxon Rank-sum Test and ANOVA were performed on each feature to select features that were most significantly correlated with CMS. It statistically tests whether the mean of the positive groups is greater than that of the negative groups or not (*P* < 0.05).

### Statistical analysis

Statistical analyses in this study were conducted using R 4.0.3. Survival curves were estimated using the Kaplan-Meier method and compared using the log-rank test, and hazard ratios were calculated using Cox proportional hazards regression. Associations between radiomic features and CMS subgroups in high-attention regions were evaluated using the Wilcoxon rank-sum test. Statistical differences between AUC values were assessed with the DeLong test. A *P* value of less than 0.05 was considered statistically significant for all tests.

## Authorship contribution

X.Z., F.G. and X.W. contributed to the study concept and design. X.Z. and X.N. contributed to data collection, analysis, draft writing and interpretation. T.W., D.C., H.X., L.Q., Y.W., Y.C., L.H., Y.Z. and Y.C. provided important advice and assistance for manuscript drafting. H.W., X.W. and E.L. contributed to data curation and annotation. Y.D., F.G. and X.W. supervised the study. All authors read and approved the final manuscript and had final responsibility for the decision to submit for publication.

## Acknowledgements

This work was funded by a grant from Shenzhen Medical Research Funds (C2303002, X.W.), a startup grant (4937084, X.W.), a direct grant (2024.175, X.W.), grants by the Faculty Postdoctoral Fellowship Scheme (FPFS/24-25/053, FPFS/23-24/061C, FPFS/23-24/060, X.W), and Research Committee – Group Research Scheme 2022-23 (WW/rc/grs2223/0560/23en, X.W.), from the Chinese University of Hong Kong, and grants from the Research Grants Council (AoE/M-401/20, R4007-23, C4024-22GF, 14104223, 11103921, and 14111522), and Health and Medical Research Fund (08192166). This work was also partially sponsored by Jiangxi Overseas High-Level Talent Project (20232BCJ25029) awarded to X.W.

## Data availability

Deidentified final results supporting this study are available for research purposes upon a reasonable written request to the corresponding author. Access to such data is available from the date of publication and requires a Data Access Agreement, which is examined and approved by the ethics committees that approved this research. Details of the analyses used in this study are fully described in the method section. The source code can be shared upon a reasonable written request to the corresponding author and requires a data use agreement.

