## Supplemental figures for "CT4CMS: Preoperative Computed Tomography-Based Consensus Molecular Subtyping Prediction in Colorectal Cancer Using Interpretable Deep Learning"

### Supplementary

**A** SYSU-CRC

| Variable | Mean (IQR) / N (%) | Univariable Cox regression HR (95% CI) | Univariable *P* | Multivariable Cox regression HR (95% CI) | Multivariable *P* |
|---|---|---|---|---|---|
| Age | 58.3 (51.0−69.0) | 1.02 (1.00−1.03) | 0.041 | 1.04 (1.02−1.06) | <0.001 |
| Sex | | | | | |
| Female (Reference) | 92 (42.6%) | | | | |
| Male | 124 (57.4%) | 1.91 (1.23−2.98) | 0.004 | 1.73 (1.09−2.73) | 0.019 |
| Grade | | | | | |
| G1 (Reference) | 65 (30.1%) | | | | |
| G2 | 136 (63.0%) | 1.56 (0.96−2.54) | 0.075 | 0.78 (0.47−1.31) | 0.354 |
| G3 | 11 (5.1%) | 2.69 (1.15−6.31) | 0.023 | 1.59 (0.64−3.94) | 0.321 |
| G4 | 4 (1.9%) | 3.21 (0.96−10.76) | 0.058 | 1.02 (0.29−3.57) | 0.977 |
| TNM stage | | | | | |
| I (Reference) | 25 (11.6%) | | | | |
| II | 75 (34.7%) | 1.42 (0.48−4.22) | 0.530 | 1.17 (0.39−3.51) | 0.785 |
| III | 62 (28.7%) | 2.54 (0.88−7.35) | 0.085 | 2.39 (0.81−7.05) | 0.113 |
| IV | 54 (25.0%) | 17.19 (6.10−48.46) | <0.001 | 18.30 (6.23−53.73) | <0.001 |
| MMR status | | | | | |
| MSI (Reference) | 20 (9.3%) | | | | |
| MSS | 196 (90.7%) | 3.69 (1.17−11.65) | 0.026 | 2.25 (0.67−7.58) | 0.190 |
| Adjuvant chemotherapy | | | | | |
| No (Reference) | 124 (57.4%) | | | | |
| Yes | 92 (42.6%) | 0.85 (0.56−1.29) | 0.441 | | |
| CMS subtype | | | | | |
| Non−CMS4 (Reference) | 155 (71.8%) | | | | |
| CMS4 | 61 (28.2%) | 2.29 (1.51−3.46) | <0.001 | 1.86 (1.20−2.88) | 0.006 |

**Figure S1. Univariable and multivariable Cox analysis of clinical factors and CMS4 subtype in the SYSU-CRC cohort.** Univariable and multivariable Cox proportional hazards analyses were performed to evaluate associations between clinical variables, CT4CMS-predicted molecular subtype, and disease-free survival (DFS) in the SYSU-CRC cohort. The CMS4 subtype was identified as a significant risk factor for poorer DFS in both univariable (*HR* = 2.29, 95% CI 1.51-3.46, *P* < 0.001) and multivariable (*HR* = 1.86, 95% CI 1.20-2.88, *P* = 0.006) analyses, indicating that CMS4 status independently predicts adverse prognosis.

**A** SYSU-CRC Stage II and III

| Variable | Mean (IQR) / N (%) | Univariable Cox regression HR (95% CI) | Univariable *P* | Multivariable Cox regression HR (95% CI) | Multivariable *P* |
|---|---|---|---|---|---|
| Age | 59.7 (52.0−69.0) | 1.06 (1.03−1.09) | <0.001 | 1.06 (1.03−1.10) | <0.001 |
| Sex | | | | | |
| Female (Reference) | 58 (42.3%) | | | | |
| Male | 79 (57.7%) | 2.83 (1.34−5.94) | 0.006 | 2.96 (1.40−6.27) | 0.005 |
| Grade | | | | | |
| G1 (Reference) | 41 (29.9%) | | | | |
| G2 | 88 (64.2%) | 1.19 (0.59−2.41) | 0.627 | | |
| G3 | 6 (4.4%) | 1.38 (0.30−6.21) | 0.679 | | |
| G4 | 2 (1.5%) | 2.03 (0.26−15.74) | 0.499 | | |
| TNM stage | | | | | |
| II (Reference) | 75 (54.7%) | | | | |
| III | 62 (45.3%) | 1.77 (0.95−3.32) | 0.074 | | |
| MMR status | | | | | |
| MSI (Reference) | 18 (13.1%) | | | | |
| MSS | 119 (86.9%) | 7.00 (0.96−50.98) | 0.055 | | |
| Adjuvant chemotherapy | | | | | |
| No (Reference) | 69 (50.4%) | | | | |
| Yes | 68 (49.6%) | 0.67 (0.36−1.27) | 0.222 | | |
| CMS subtype | | | | | |
| Non−CMS4 (Reference) | 102 (74.5%) | | | | |
| CMS4 | 35 (25.5%) | 2.26 (1.20−4.25) | 0.012 | 1.95 (1.03−3.68) | 0.040 |

**Figure S2. Univariable and multivariable Cox analysis of clinical factors and CMS4 subtype in stage II and III colorectal cancer within the SYSU-CRC cohort.** Univariable and multivariable Cox proportional hazards analyses were performed to evaluate associations between clinical variables, CT4CMS-predicted molecular subtype, and disease-free survival (DFS) in patients with stage II-III colorectal cancer. The CMS4 subtype was identified as a significant risk factor for poorer DFS in both univariable (*HR* = 2.26, 95% CI 1.20-4.25, *P* = 0.012) and multivariable (*HR* = 1.95, 95% CI 1.03-3.68, *P* = 0.040) analyses, indicating that CMS4 status independently predicts adverse prognosis.

**A** SYSU-SAH

| Variable | Mean (IQR) / N (%) | Univariable Cox regression HR (95% CI) | Univariable *P* | Multivariable Cox regression HR (95% CI) | Multivariable *P* |
|---|---|---|---|---|---|
| Age | 59.8 (51.0−68.0) | 1.01 (1.00−1.02) | 0.105 | | |
| Sex | | | | | |
| Female (Reference) | 439 (40.0%) | | | | |
| Male | 658 (60.0%) | 1.71 (1.27−2.31) | <0.001 | 1.68 (1.24−2.27) | 0.001 |
| Grade | | | | | |
| G1 (Reference) | 202 (18.4%) | | | | |
| G2 | 860 (78.4%) | 1.20 (0.84−1.72) | 0.325 | 1.08 (0.75−1.56) | 0.681 |
| G3 | 27 (2.5%) | 2.47 (1.23−4.97) | 0.011 | 1.37 (0.67−2.80) | 0.395 |
| G4 | 8 (0.7%) | 3.76 (1.34−10.56) | 0.012 | 1.29 (0.45−3.71) | 0.631 |
| TNM stage | | | | | |
| I (Reference) | 30 (2.7%) | | | | |
| II | 919 (83.8%) | 1.24 (0.45−3.36) | 0.679 | 1.06 (0.38−2.93) | 0.910 |
| III | 87 (7.9%) | 2.93 (1.04−8.29) | 0.043 | 2.55 (0.89−7.34) | 0.082 |
| IV | 61 (5.6%) | 26.56 (9.53−74.04) | <0.001 | 20.32 (7.14−57.81) | <0.001 |
| MMR status | | | | | |
| MSI (Reference) | 111 (10.1%) | | | | |
| MSS | 986 (89.9%) | 1.80 (1.07−3.04) | 0.026 | 1.67 (0.97−2.86) | 0.064 |
| Adjuvant chemotherapy | | | | | |
| No (Reference) | 658 (60.0%) | | | | |
| Yes | 439 (40.0%) | 1.05 (0.79−1.38) | 0.738 | | |
| CMS subtype | | | | | |
| Non−CMS4 (Reference) | 814 (74.2%) | | | | |
| CMS4 | 283 (25.8%) | 2.03 (1.53−2.68) | <0.001 | 1.78 (1.34−2.36) | <0.001 |

Univariable axis: 1 2.7 7.4 20.1 54.6; Multivariable axis: 0.4 1 2.7 7.4 20.1 54.6

**Figure S3. Univariable and multivariable Cox analysis of clinical factors and CMS4 subtype in the SYSU-SAH cohort.** Univariable and multivariable Cox proportional hazards analyses were conducted to assess the associations of clinical variables and CT4CMS-predicted molecular subtype with disease-free survival (DFS) in the SYSU-SAH cohort. The CMS4 subtype was significantly associated with worse DFS in both univariable ($HR$ = 2.03, 95% CI 1.53-2.68, $P$ < 0.001) and multivariable ($HR$ = 1.78, 95% CI 1.34-2.36, $P$ < 0.001) analyses, confirming CMS4 as an independent prognostic risk factor.

**A** SYSU-SAH Stage II and III

| Variable | Mean (IQR) / N (%) | Univariable Cox regression HR (95% CI) | Univariable Cox regression *P* | Multivariable Cox regression HR (95% CI) | Multivariable Cox regression *P* |
|---|---|---|---|---|---|
| Age | 60.2 (52.0–69.0) | 1.02 (1.01–1.04) | 0.001 | 1.03 (1.01–1.04) | <0.001 |
| Sex | | | | | |
| Female (Reference) | 400 (39.8%) | | | | |
| Male | 606 (60.2%) | 1.83 (1.27–2.62) | 0.001 | 1.83 (1.27–2.63) | 0.001 |
| Grade | | | | | |
| G1 (Reference) | 171 (17.0%) | | | | |
| G2 | 808 (80.3%) | 1.41 (0.90–2.22) | 0.136 | | |
| G3 | 21 (2.1%) | 1.50 (0.52–4.33) | 0.457 | | |
| G4 | 6 (0.6%) | 3.05 (0.72–12.95) | 0.131 | | |
| TNM stage | | | | | |
| II (Reference) | 919 (91.4%) | | | | |
| III | 87 (8.6%) | 2.29 (1.53–3.44) | <0.001 | 2.69 (1.78–4.08) | <0.001 |
| MMR status | | | | | |
| MSI (Reference) | 105 (10.4%) | | | | |
| MSS | 901 (89.6%) | 1.92 (1.05–3.51) | 0.034 | 1.87 (1.01–3.48) | 0.047 |
| Adjuvant chemotherapy | | | | | |
| No (Reference) | 595 (59.1%) | | | | |
| Yes | 411 (40.9%) | 0.99 (0.71–1.37) | 0.937 | | |
| CMS subtype | | | | | |
| Non–CMS4 (Reference) | 758 (75.3%) | | | | |
| CMS4 | 248 (24.7%) | 1.74 (1.24–2.44) | 0.001 | 1.72 (1.22–2.41) | 0.002 |

**Figure S4. Univariable and multivariable Cox analysis of clinical factors and CMS4 subtype in stage II and III colorectal cancer within the SYSU-SAH cohort.** Univariable and multivariable Cox proportional hazards analyses were conducted to assess the associations of clinical variables and CT4CMS-predicted molecular subtype with disease-free survival (DFS) in patients with stage II-III colorectal cancer. The CMS4 subtype was significantly associated with worse DFS in both univariable ($HR = 1.74$, 95% CI 1.24-2.44, $P = 0.001$) and multivariable ($HR = 1.72$, 95% CI 1.22-2.41, $P = 0.002$) analyses, confirming CMS4 as an independent prognostic risk factor.

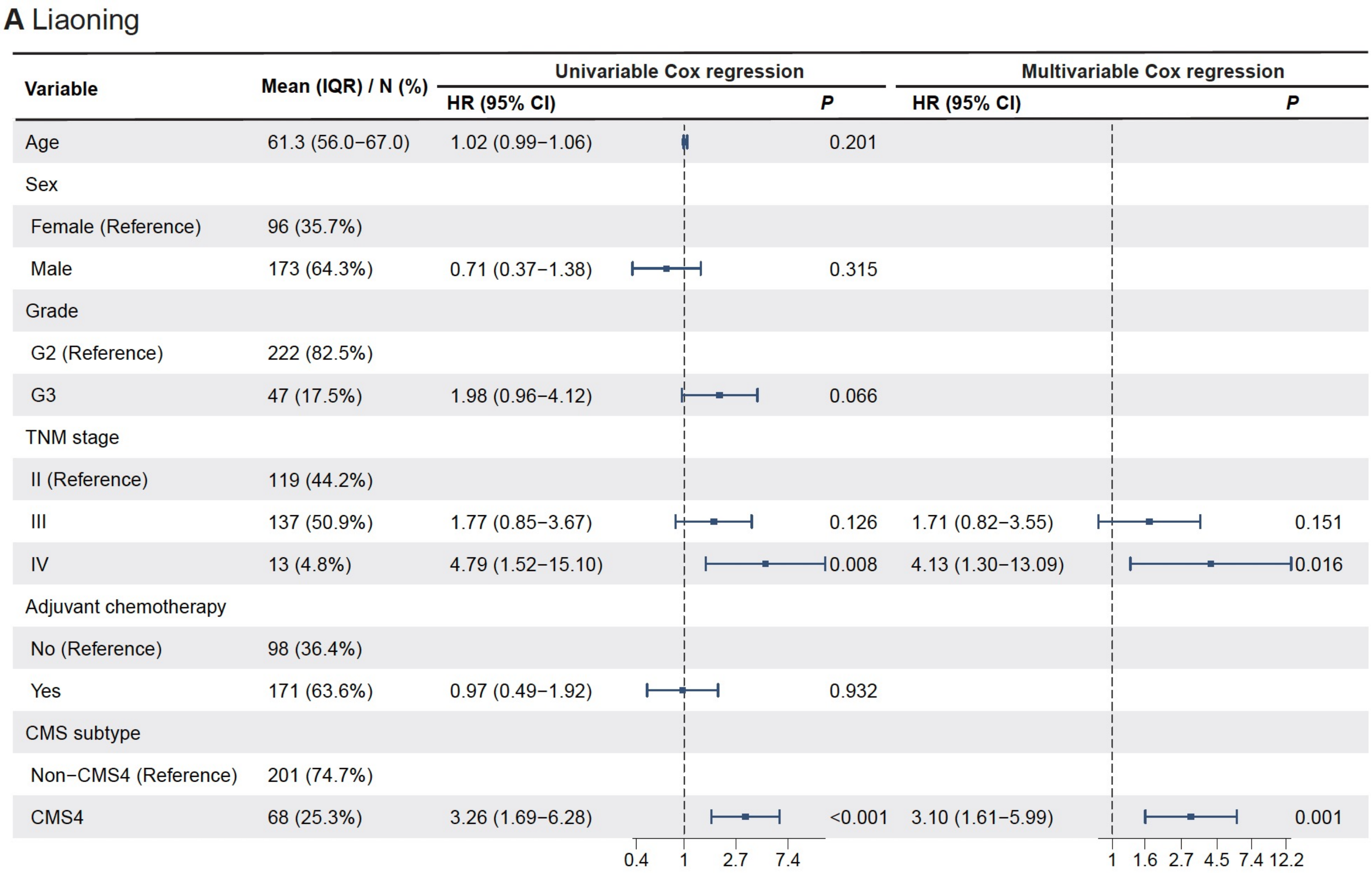

**A** Liaoning

| Variable | Mean (IQR) / N (%) | Univariable Cox regression HR (95% CI) | P | Multivariable Cox regression HR (95% CI) | P |
|---|---|---|---|---|---|
| Age | 61.3 (56.0−67.0) | 1.02 (0.99−1.06) | 0.201 | | |
| Sex | | | | | |
| Female (Reference) | 96 (35.7%) | | | | |
| Male | 173 (64.3%) | 0.71 (0.37−1.38) | 0.315 | | |
| Grade | | | | | |
| G2 (Reference) | 222 (82.5%) | | | | |
| G3 | 47 (17.5%) | 1.98 (0.96−4.12) | 0.066 | | |
| TNM stage | | | | | |
| II (Reference) | 119 (44.2%) | | | | |
| III | 137 (50.9%) | 1.77 (0.85−3.67) | 0.126 | 1.71 (0.82−3.55) | 0.151 |
| IV | 13 (4.8%) | 4.79 (1.52−15.10) | 0.008 | 4.13 (1.30−13.09) | 0.016 |
| Adjuvant chemotherapy | | | | | |
| No (Reference) | 98 (36.4%) | | | | |
| Yes | 171 (63.6%) | 0.97 (0.49−1.92) | 0.932 | | |
| CMS subtype | | | | | |
| Non−CMS4 (Reference) | 201 (74.7%) | | | | |
| CMS4 | 68 (25.3%) | 3.26 (1.69−6.28) | <0.001 | 3.10 (1.61−5.99) | 0.001 |



**Figure S5. Univariable and multivariable Cox analysis of clinical factors and CMS4 subtype in the Liaoning cohort.** Univariable and multivariable Cox proportional hazards analyses were performed to assess associations of clinical variables and CT4CMS-predicted molecular subtype with disease-free survival (DFS) in the external Liaoning cohort. The CMS4 subtype was consistently associated with an increased risk of disease recurrence in both univariable ($HR = 3.26$, 95% CI 1.69-6.28, $P < 0.001$) and multivariable ($HR = 3.10$, 95% CI 1.61-5.99, $P = 0.001$) analyses, confirming CMS4 as an independent adverse prognostic factor.

**A** Liaoning Stage II and III

| Variable | Mean (IQR) / N (%) | Univariable Cox regression HR (95% CI) | Univariable Cox regression *P* | Multivariable Cox regression HR (95% CI) | Multivariable Cox regression *P* |
|---|---|---|---|---|---|
| Age | 61.3 (56.0−67.2) | 1.03 (0.99−1.07) | 0.200 | | |
| Sex | | | | | |
| Female (Reference) | 89 (34.8%) | | | | |
| Male | 167 (65.2%) | 0.79 (0.39−1.60) | 0.514 | | |
| Grade | | | | | |
| G2 (Reference) | 213 (83.2%) | | | | |
| G3 | 43 (16.8%) | 1.54 (0.66−3.56) | 0.315 | | |
| TNM stage | | | | | |
| II (Reference) | 119 (46.5%) | | | | |
| III | 137 (53.5%) | 1.76 (0.85−3.66) | 0.129 | | |
| Adjuvant chemotherapy | | | | | |
| No (Reference) | 93 (36.3%) | | | | |
| Yes | 163 (63.7%) | 0.93 (0.45−1.90) | 0.844 | | |
| CMS subtype | | | | | |
| Non−CMS4 (Reference) | 192 (75.0%) | | | | |
| CMS4 | 64 (25.0%) | 3.34 (1.66−6.69) | 0.001 | 3.34 (1.66−6.69) | 0.001 |

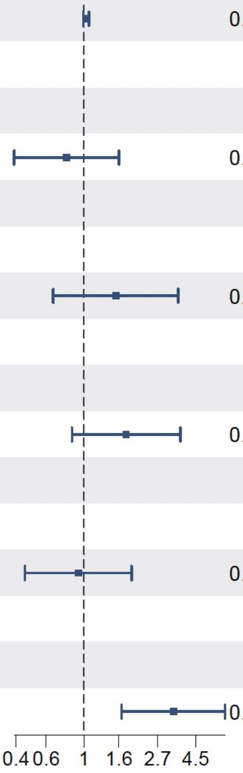


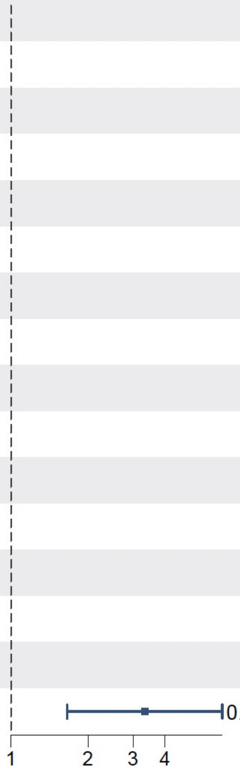


**Figure S6. Univariable and multivariable Cox analysis of clinical factors and CMS4 subtype in stage II and III colorectal cancer within the Liaoning cohort.** Univariable and multivariable Cox proportional hazards analyses were performed to assess associations of clinical variables and CT4CMS-predicted molecular subtype with disease-free survival (DFS) in patients with stage II-III colorectal cancer. The CMS4 subtype was consistently associated with an increased risk of disease recurrence in both univariable and multivariable ($HR = 3.10$, 95% CI 1.61-5.99, $P = 0.001$) analyses, confirming CMS4 as an independent adverse prognostic factor.

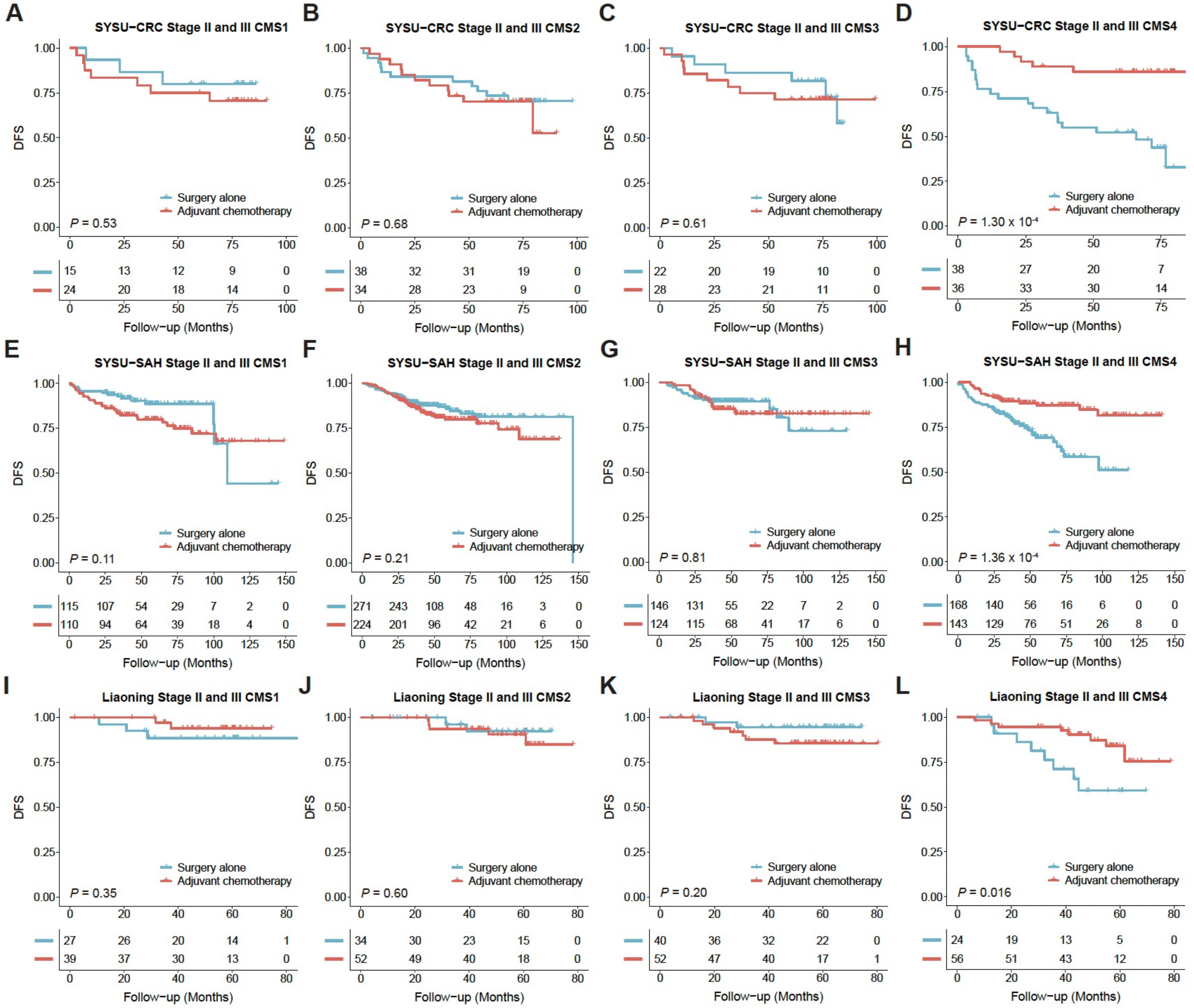


**Figure S7. Adjuvant chemotherapy benefit across CMS subtypes in stage II and III colorectal cancer. (A-L)** Kaplan-Meier curves of disease-free survival (DFS) comparing patients receiving surgery alone versus adjuvant chemotherapy within each consensus molecular subtype (CMS) across the SYSU-CRC **(A-D)**, SYSU-SAH **(E-H)**, and Liaoning **(I-L)** cohorts. Among all subtypes, CMS4 patients derived a significant survival benefit from adjuvant chemotherapy compared with surgery alone in the SYSU-CRC ($P = 1.30 \times 10^{-4}$), SYSU-SAH ($P = 1.36 \times 10^{-4}$) and Liaoning ($P = 0.016$) cohorts, whereas the other CMS groups showed no statistically significant differences.

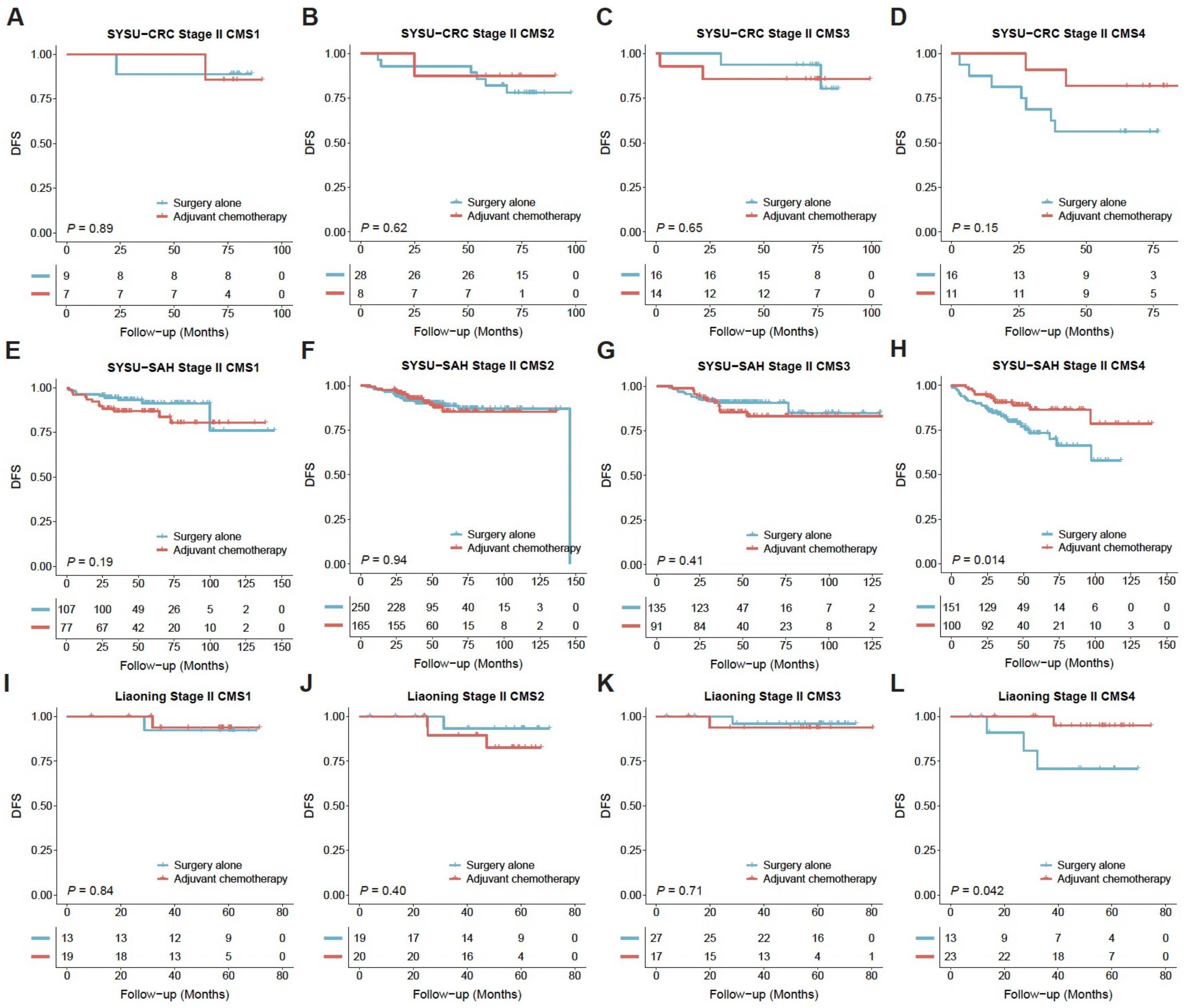


**Figure S8. Adjuvant chemotherapy benefit across CMS subtypes in stage II colorectal cancer. (A-L)** Kaplan-Meier curves of disease-free survival (DFS) comparing patients receiving surgery alone versus adjuvant chemotherapy within each consensus molecular subtype (CMS) across the SYSU-CRC **(A-D)**, SYSU-SAH **(E-H)**, and Liaoning **(I-L)** cohorts. Among all subtypes, CMS4 patients showed the clearest trend toward improved survival with adjuvant chemotherapy compared with surgery alone, reaching statistical significance in the SYSU-SAH ($P$ = 0.014) and Liaoning ($P$ = 0.042) cohorts. In the SYSU-CRC cohort, CMS4 patients also exhibited a favorable trend, whereas the other CMS groups showed no significant differences between treatment arms.

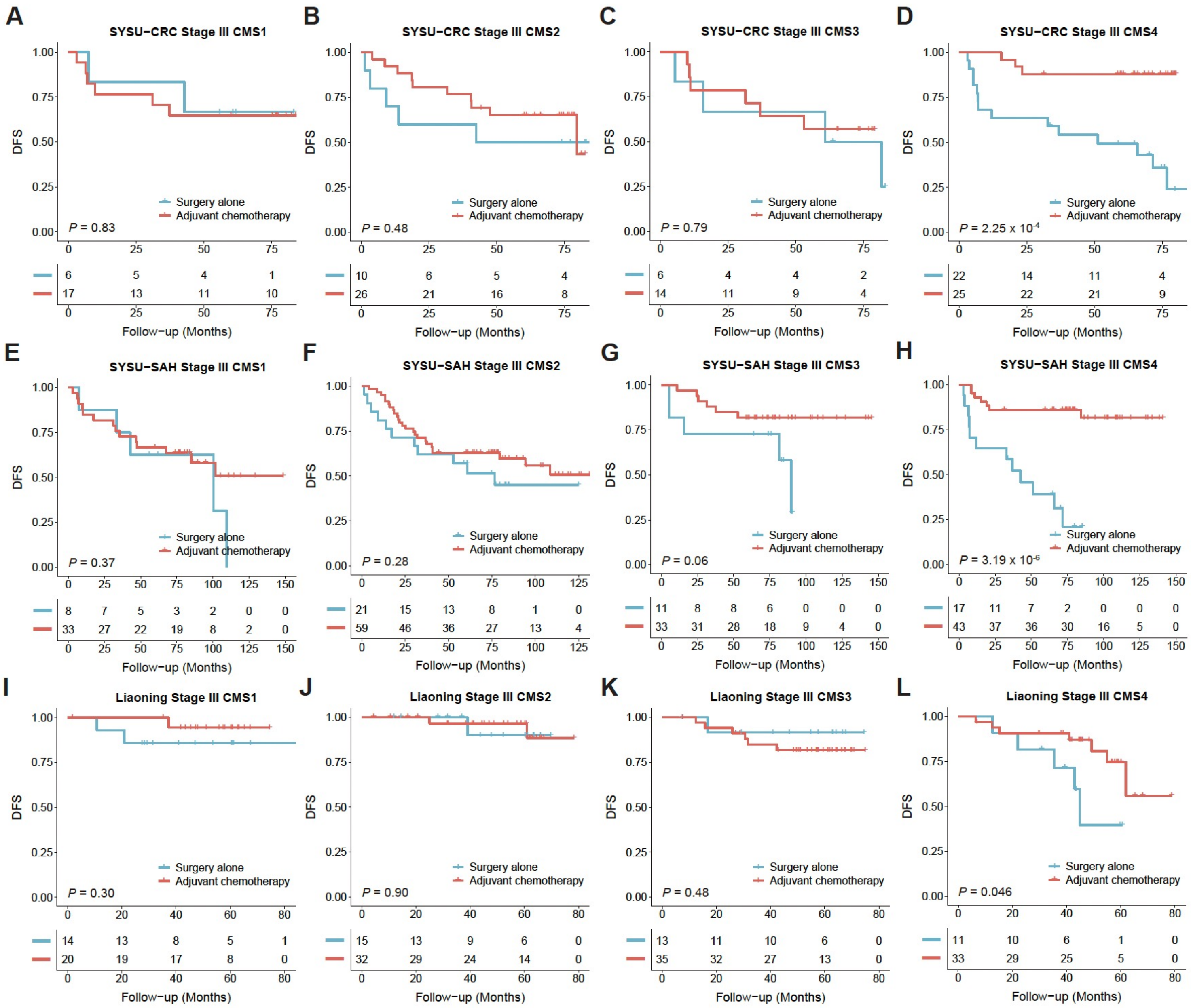


**Figure S9. Adjuvant chemotherapy benefit across CMS subtypes in stage III colorectal cancer. (A-L)** Kaplan-Meier curves of disease-free survival (DFS) comparing patients receiving surgery alone versus adjuvant chemotherapy within each consensus molecular subtype (CMS) across the SYSU-CRC **(A-D)**, SYSU-SAH **(E-H)**, and Liaoning **(I-L)** cohorts. Among all subtypes, CMS4 patients derived a significant survival benefit from adjuvant chemotherapy compared with surgery alone in the SYSU-CRC ($P = 2.25 \times 10^{-4}$), SYSU-SAH ($P = 3.19 \times 10^{-6}$) and Liaoning ($P = 0.046$). In contrast, the other CMS groups showed no statistically significant differences.

**A** SYSU-CRC Stage II and III

| Subtype | Adjuvant chemotherapy (Yes / No) | HR (95% CI) | *P* |
|---|---|---|---|
| CMS1 | 24/15 | 1.54 (0.40−5.97) | 0.527 |
| CMS2 | 34/38 | 1.19 (0.52−2.77) | 0.677 |
| CMS3 | 28/22 | 1.32 (0.45−3.91) | 0.613 |
| CMS4 | 36/38 | 0.18 (0.07−0.48) | <0.001 |

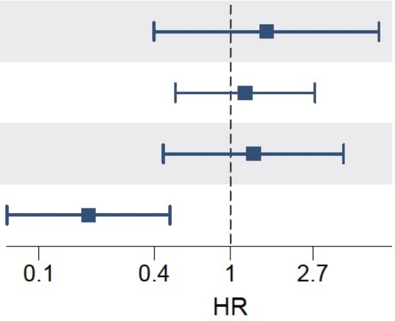


**B** SYSU-SAH Stage II and III

| Subtype | Adjuvant chemotherapy (Yes / No) | HR (95% CI) | *P* |
|---|---|---|---|
| CMS1 | 110/115 | 1.69 (0.88−3.27) | 0.112 |
| CMS2 | 224/271 | 1.33 (0.85−2.09) | 0.206 |
| CMS3 | 124/146 | 1.08 (0.57−2.07) | 0.814 |
| CMS4 | 143/168 | 0.36 (0.21−0.62) | <0.001 |

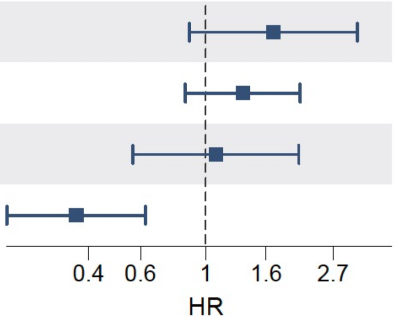


**C** Liaoning Stage II and III

| Subtype | Adjuvant chemotherapy (Yes / No) | HR (95% CI) | *P* |
|---|---|---|---|
| CMS1 | 39/27 | 0.44 (0.07−2.62) | 0.351 |
| CMS2 | 52/34 | 1.55 (0.30−8.01) | 0.596 |
| CMS3 | 52/40 | 2.69 (0.56−12.96) | 0.198 |
| CMS4 | 56/24 | 0.31 (0.12−0.85) | 0.016 |

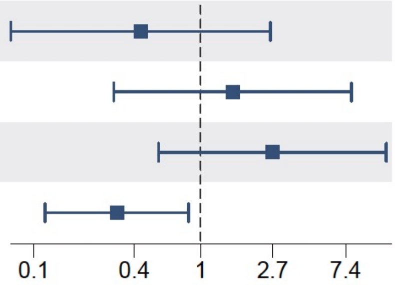


**Figure S10. Univariable Cox analysis of adjuvant chemotherapy benefit within CMS subtypes in stage II and III colorectal cancer. (A-C)** Forest plots showing univariable Cox proportional hazards analysis comparing adjuvant chemotherapy versus surgery alone for disease-free survival (DFS) in each consensus molecular subtype (CMS) across the SYSU-CRC **(A)**, SYSU-SAH **(B)**, and Liaoning **(C)** cohorts. The CMS4 subgroup consistently demonstrated a significant benefit from adjuvant chemotherapy with hazard ratios (HRs) below 1.0 across all datasets. In contrast, no significant improvement in DFS was observed with adjuvant chemotherapy among CMS1-3 subtypes.

**A** SYSU-CRC Stage II

| Subtype | Adjuvant chemotherapy (Yes / No) | HR (95% CI) | *P* |
|---|---|---|---|
| CMS1 | 7/9 | 1.21 (0.08−19.40) | 0.892 |
| CMS2 | 8/28 | 0.59 (0.07−4.90) | 0.620 |
| CMS3 | 14/16 | 1.59 (0.21−11.83) | 0.650 |
| CMS4 | 11/16 | 0.33 (0.07−1.59) | 0.147 |

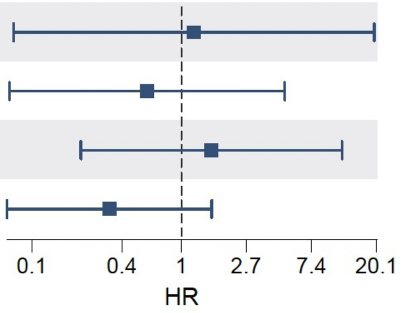


**B** SYSU-SAH Stage II

| Subtype | Adjuvant chemotherapy (Yes / No) | HR (95% CI) | *P* |
|---|---|---|---|
| CMS1 | 77/107 | 1.76 (0.74−4.19) | 0.195 |
| CMS2 | 165/250 | 0.98 (0.52−1.83) | 0.944 |
| CMS3 | 91/135 | 1.38 (0.64−2.98) | 0.407 |
| CMS4 | 100/151 | 0.45 (0.23−0.86) | 0.014 |

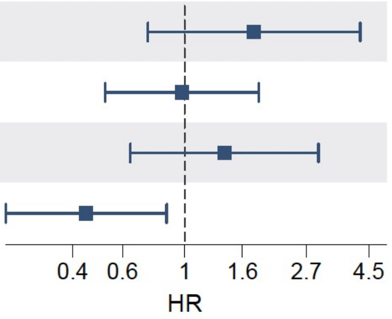


**C** Liaoning Stage II

| Subtype | Adjuvant chemotherapy (Yes / No) | HR (95% CI) | *P* |
|---|---|---|---|
| CMS1 | 19/13 | 0.76 (0.05−12.11) | 0.844 |
| CMS2 | 20/19 | 2.57 (0.27−24.72) | 0.397 |
| CMS3 | 17/27 | 1.67 (0.10−26.75) | 0.714 |
| CMS4 | 23/13 | 0.14 (0.01−1.30) | 0.042 |

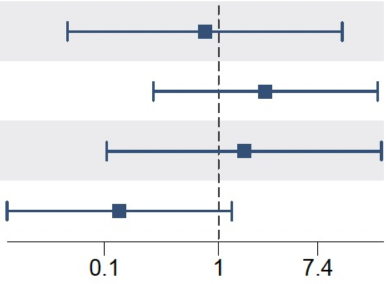


**Figure S11. Univariable Cox analysis of adjuvant chemotherapy benefit within CMS subtypes in stage II colorectal cancer. (A-C)** Forest plots showing univariable Cox proportional hazards analysis comparing adjuvant chemotherapy versus surgery alone for disease-free survival (DFS) in each consensus molecular subtype (CMS) across the SYSU-CRC **(A)**, SYSU-SAH **(B)**, and Liaoning **(C)** cohorts. The CMS4 subgroup consistently demonstrated a favorable association between adjuvant chemotherapy and improved DFS across all datasets. No significant chemotherapy benefit was observed across the three independent cohorts in the CMS1-3 subtypes.

**A** SYSU-CRC Stage III

| Subtype | Adjuvant chemotherapy (Yes / No) | HR (95% CI) | *P* |
|---|---|---|---|
| CMS1 | 17/6 | 1.19 (0.24−5.88) | 0.834 |
| CMS2 | 26/10 | 0.68 (0.23−2.00) | 0.476 |
| CMS3 | 14/6 | 0.83 (0.21−3.33) | 0.792 |
| CMS4 | 25/22 | 0.13 (0.04−0.47) | <0.001 |

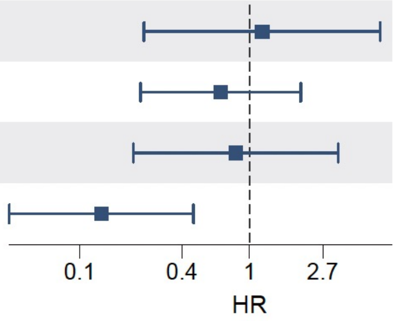


**B** SYSU-SAH Stage III

| Subtype | Adjuvant chemotherapy (Yes / No) | HR (95% CI) | *P* |
|---|---|---|---|
| CMS1 | 33/8 | 0.63 (0.23−1.76) | 0.372 |
| CMS2 | 59/21 | 0.68 (0.33−1.38) | 0.282 |
| CMS3 | 33/11 | 0.33 (0.10−1.08) | 0.054 |
| CMS4 | 43/17 | 0.14 (0.05−0.36) | <0.001 |

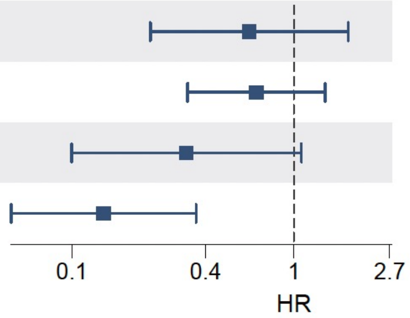


**C** Liaoning Stage III

| Subtype | Adjuvant chemotherapy (Yes / No) | HR (95% CI) | *P* |
|---|---|---|---|
| CMS1 | 20/14 | 0.30 (0.03−3.37) | 0.302 |
| CMS2 | 32/15 | 0.86 (0.08−9.54) | 0.905 |
| CMS3 | 35/13 | 2.12 (0.25−17.59) | 0.477 |
| CMS4 | 33/11 | 0.31 (0.09−1.04) | 0.046 |

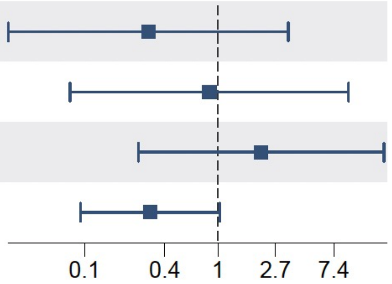


**Figure S12. Univariable Cox analysis of adjuvant chemotherapy benefit within CMS subtypes in stage III colorectal cancer. (A-C)** Forest plots showing univariable Cox proportional hazards analysis comparing adjuvant chemotherapy versus surgery alone for disease-free survival (DFS) in each consensus molecular subtype (CMS) across the SYSU-CRC **(A)**, SYSU-SAH **(B)**, and Liaoning **(C)** cohorts. The CMS4 subgroup consistently demonstrated a significant benefit from adjuvant chemotherapy and improved DFS. In contrast, no significant survival benefit was observed for CMS1–3 subtypes across the three independent cohorts.

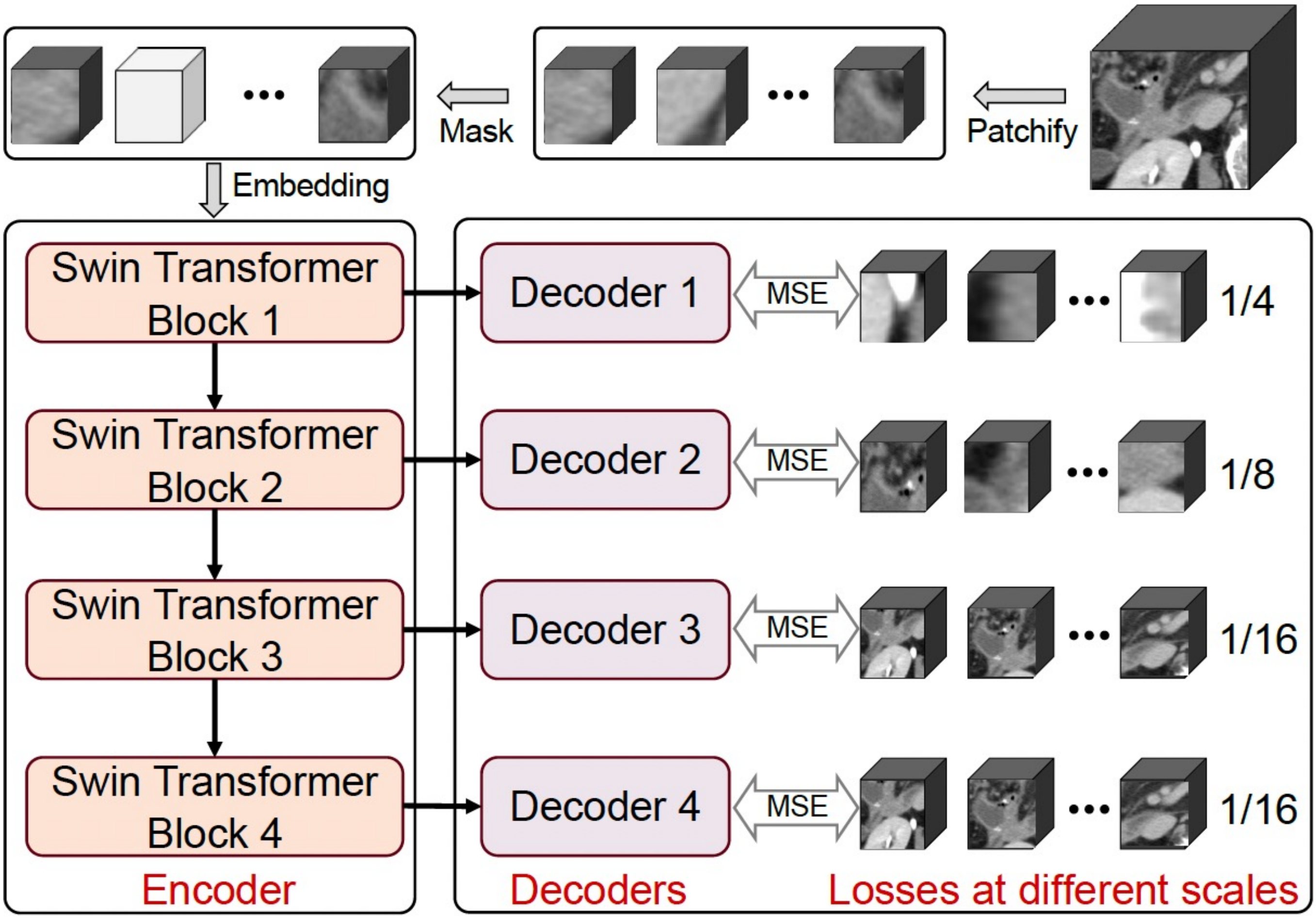


**Figure S13. Self-supervised pretraining of the multi-scale backbone in CT4CMS.** Schematic illustration of the multi-scale self-supervised pretraining framework used for the CT4CMS backbone. Three-dimensional CT volumes were first divided into patches and randomly masked. The visible patches were embedded and passed through a hierarchical Swin-Transformer encoder consisting of four successive blocks (Blocks 1-4). Each encoder block was linked to a corresponding decoder that reconstructed the masked patches at multiple spatial resolutions (1/4, 1/8, 1/16 of the original scale). Mean-squared-error (MSE) losses were computed at each decoding level to guide multi-scale feature learning, enabling the backbone to capture both local and global contextual representations from unlabeled CT data.

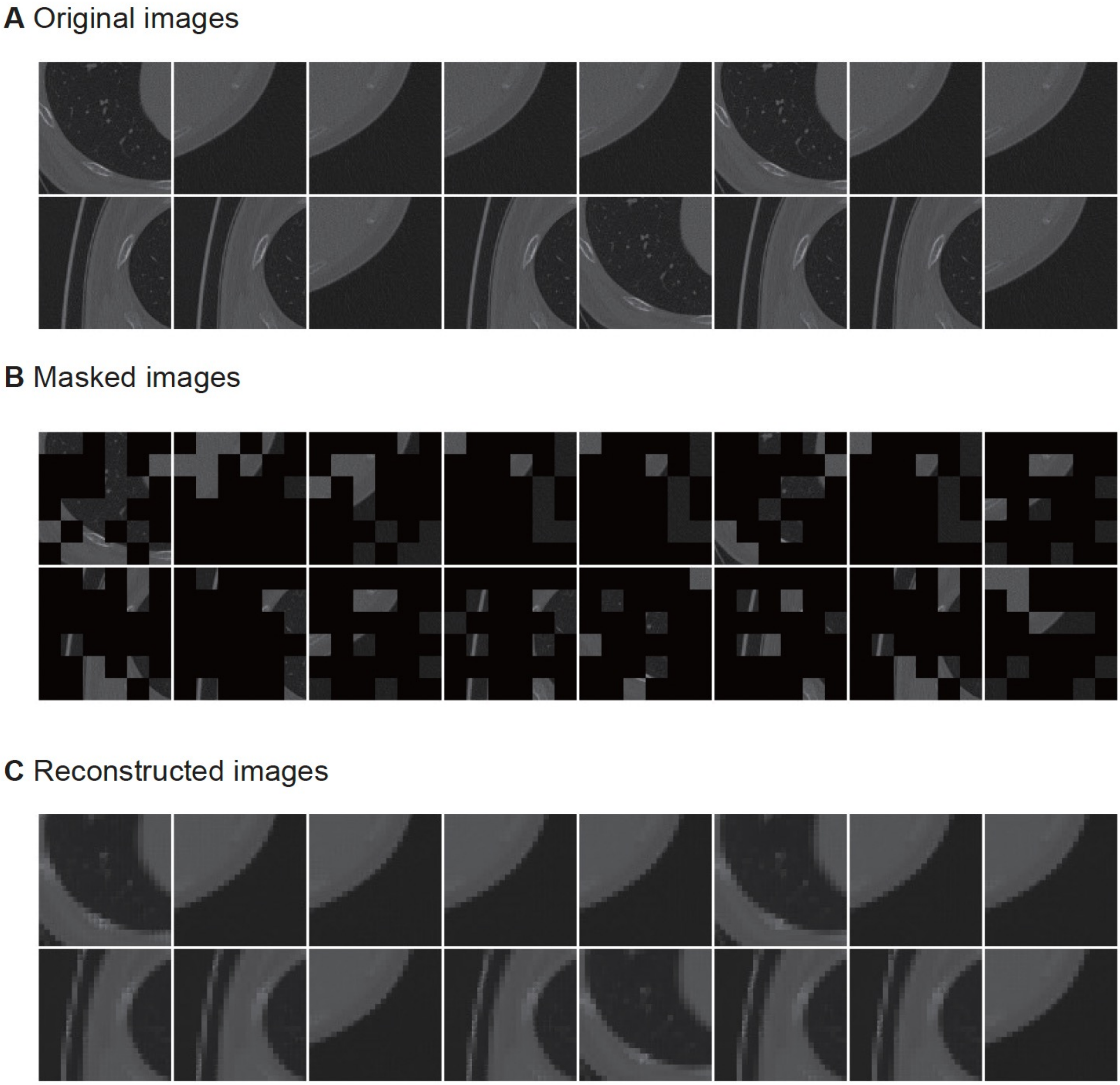


**Figure S14. Reconstruction results of the multiscale masked image modeling (MIM) approach with a mask ratio of 75%.** The three rows show the original images **(A)**, the masked images **(B)**, and the reconstructed images **(C)**, respectively. The results demonstrated that multiscale MIM could restore lost information from the randomly masked context.
